# Factors associated with effectiveness of interventions to prevent obesity in children: a synthesis of evidence from 204 randomized trials

**DOI:** 10.1101/2024.06.19.24309160

**Authors:** Annabel L Davies, Francesca Spiga, Deborah M Caldwell, Jelena Savović, Jennifer C Palmer, Eve Tomlinson, Theresa HM Moore, Carolyn D Summerbell, Julian PT Higgins

**Affiliations:** Population Health Sciences, Bristol Medical School, University of Bristol, Bristol, UK; NIHR Applied Research Collaboration West (ARC West) at University Hospitals Bristol and Weston NHS Foundation Trust, Bristol, UK; Bristol Medical School, University of Bristol, Bristol, UK; Department of Sport and Exercise Sciences, Durham University, Durham, UK; Fuse - Centre for Translational Research in Public Health, Newcastle upon Tyne, UK

**Keywords:** Obesity, Physical activity, Diet, Behaviour change, Evidence synthesis, Effectiveness

## Abstract

**Objective:** To identify effective characteristics of behaviour change (physical activity and diet) interventions that prevent obesity in children aged 5 to 18 years.

**Design:** A Bayesian multi-level meta-regression analysis of randomized trial results, with intervention and trial characteristics coded according to an analytic framework co-developed with stakeholders.

**Data source:** Two Cochrane systematic reviews of the effects of interventions to prevent obesity in children, 5 to 11 years and 12 to 18 years, both updated in 2024.

**Main outcome measures:** Mean difference (MD) in change from baseline in age- and sex-standardized BMI measured as a Z-score (zBMI). Results that had been reported as (unstandardized) BMI or BMI percentile were converted to zBMI using bespoke mapping techniques.

**Results:** We included 204 trials (255 intervention arms) reporting data on at least one of the main outcome scales. Interventions were effective on average (MD in zBMI −0.037, 95% credible interval −0.053 to −0.022). The greatest effects were associated with medium term follow-up (9 to <15 months) and older children (12 to 18 years). We found evidence of small but beneficial effects for interventions targeting physical activity alone compared with diet alone (difference in MDs −0.227, −0.362 to −0.090) and small unfavorable effects for interventions that involved a change to the structural environment (the majority of changes were in the school food environment) (difference in MDs 0.05, 0.017 to 0.085). Accounting for interactions between covariates, we found that the most effective combination of intervention characteristics was to intervene in the school setting, with an individualized element to delivery, targeting physical activity, using multiple strategies of short duration and high intensity, and involving modification of behaviour through participation in activities.

**Conclusions:** The most effective characteristic to include in a behaviour change intervention to prevent obesity in children aged 5-18 years was targeting of physical activity. This should not be interpreted as evidence that attempts to modify diet are not beneficial. Being physically active and consuming a healthy diet during childhood offer many important benefits beyond contributing to healthy weight and growth. Our findings suggest that interventions to prevent obesity in children should consider focusing primarily on the promotion of physical activity and consider other effective characteristics we identify here.

**Key messages:** *What is already known on this topic:*

- Rising population levels of childhood overweight and obesity present a global challenge.
- Many interventions have been developed and evaluated to try and prevent obesity in children and young people.
- The most effective characteristics of these interventions are not well understood.

*What this study adds:*

- This re-analysis of the results of 204 randomized trials of diverse interventions seeks to identify effective characteristics of behaviour change (physical activity and diet) interventions.
- The most effective characteristic to include in a behaviour change intervention may be targeting physical activity.
- Other useful features of interventions appear to be individualized delivery, using multiple strategies, being intense and of short duration, and involving participation in activities.

## 1 Introduction

Rising population levels of childhood overweight and obesity present a global challenge [1], with profound implications for public health and health services [2]. Children and adolescents living with obesity are more likely to experience reduced health-related quality of life and, for adolescents, a number of comorbidities [3]. It is therefore important to prevent childhood obesity, both to ensure good long-term physical and mental health and to help children realise their full potential [4]. Prevention requires effective interventions to change behaviour in relation to dietary habits and physical activity. Such behaviour-change interventions typically contain multiple techniques, or components [5], which may act individually or in combination to determine the effectiveness of the intervention.

We recently conducted two Cochrane systematic reviews of 244 randomized controlled trials of behaviour change interventions aimed at preventing obesity in children (aged 5 to 11 years and 12 to 18 years, respectively) [6, 7]. Interventions were grouped according to whether they targeted physical activity, diet or both. While we found some modest beneficial effects, on average, of physical activity interventions, either alone or in combination with diet, there remained substantial between-trial variability in results across trials within each of the broad comparisons. This heterogeneity is most likely caused by variation in the characteristics of interventions included in each category. The characteristics varied in many different dimensions, including whether they targeted diet or physical activity or both, the degree of home (family) engagement, the degree of active participation by the children, the number of different strategies employed concurrently, the mode of delivery, the intensity and the duration.

In this paper we report results of a re-analysis of these trials using all types of BMI outcomes reported, age groups (5-18 years) and follow-up times in a single comprehensive analysis. Our aim is to identify characteristics of these behaviour change interventions that are most strongly associated with their effectiveness in preventing obesity in children. We employ an analytic framework co-produced with stakeholders [8], statistical methods for mapping different outcomes onto a single measurement scale [9], and a bespoke complex meta-regression model [10].

## 2 Methods

### 2.1 Data and previous analyses

We used data from our two recently published Cochrane systematic reviews and meta-analyses, reported in detail elsewhere [6, 7]. In brief, the reviews included randomized trials (either individually randomized or cluster randomized) of behaviour-change interventions that targeted dietary and/or physical activity in any setting that aimed to prevent obesity in children aged 5 to 18 years. Trials were excluded if they were restricted to children living with overweight and/or obesity as we were interested in prevention rather than treatment. Our outcomes of interest were unstandardized body mass index (BMI) and age- and sex-standardized BMI measured as z-scores (zBMI) or percentiles. For the Cochrane reviews we calculated mean differences, with standard errors, in change from baseline between intervention groups in each trial, where appropriate adjusting the standard errors for clustering [6, 7]. We use these mean differences and standard errors as the observations in our model. The dataset includes multi-arm trials as well as trials with multiple time points. We categorize follow up times into short- (12 weeks to <9 months), medium- (9 months to <15 months) and long-term (15 months or more).

### 2.2 Intervention-level coding

We developed an analytic framework to inform the synthesis. First, we sought to include intervention-level characteristics considered most likely to be associated with effectiveness. To compile these, we reviewed the international literature for relevant theories and frameworks. We engaged extensively with children and young people, teachers and public health professionals. We presented these groups with potential items and asked them to suggest characteristics they deemed most likely to be effective. Development of the framework is described in detail elsewhere [8].

The finalized analytic framework comprises 12 main intervention characteristics (A to L in Box 1). We coded each intervention in each trial according to these characteristics as described previously [8]. Control arms, which we define as the absence of any intervention, were not coded. To reduce the number of variables, we applied dichotomizations to categorical variables aiming for divisions that resulted in the most even split of the data. In addition, we dichotomized intervention duration into ‘long’ and ‘short’ either side of the median duration across the trials. These dichotomous variables, which we call intervention-level indicators are also provided in Box 1, with results of the coding presented in Section A of the Supplementary Material.

#### Box 1

The 12 intervention characteristics (and 25 resulting intervention-level indicators) included in our analytic framework.

A. The setting of the intervention (four sub-questions): whether the intervention…
  a. was delivered in a school
  b. was delivered in the home
  c. was delivered in the community
  d. included a home activity
B. The mode of delivery (three sub-questions): whether the intervention was delivered to the child…
  a. as part of a group of children
  b. individually
  c. electronically
C. The change of behaviour targeted by the intervention (two sub-questions):
  a. diet behaviours
  b. physical activity behaviours
D. The multi-factor nature of the intervention and its delivery (three sub-questions): whether the intervention was applied…
  a. using multiple (three or more) different strategies
  b. in a single phase
  c. continuously
E. Intensity and duration (three sub-questions): whether the intervention…
  a. was long (vs short) in total duration
  b. was long (vs short) in duration at its peak intensity
  c. involved a high (vs low) level of engagement at its peak intensity
F. Whether the intervention was integrated into usual activities
G. Whether there was flexibility in how the intervention can be implemented
H. Whether there was a level of choice available to children experiencing the intervention
I. Whether the intervention was considered to be enjoyable for the recipients (the ‘fun factor’)
J. Whether the person/people delivering the intervention were likely to resonate with (inspire) the children
K. The mechanism(s) of action employed (four sub-questions): whether the intervention had an explicit component…
  a. requiring the child to participate
  b. providing education/information to the child
  c. aiming to change the social environment of the child
  d. aiming to change the physical environment of the child
L. Whether there were commercial interests involved

### 2.3 Trial-level coding

In addition to intervention-level characteristics, it is likely that the effectiveness of interventions depends on characteristics of the participants. To investigate the possible impact of some broad societal inequities between populations, we defined trial-level indicators capturing the income status of the country (high vs non-high) and whether the trial specifically targeted participants (or communities) with low socioeconomic status (SES). Details of how these trial-level indicators were categorized can be found in our Cochrane reviews [6, 7].

Rather than conducting separate analyses for the two age groups (as set-out in our initial protocol [11]), we analysed the combined data set and included age group (5-11 versus 12-18) as another trial-level indicator. This allowed us to examine the differential effectiveness by age group, as well as to investigate how other factors interact with age, and provided additional power when examining factors across age groups.

### 2.4 Time-point-level coding

We coded follow-up time as a categorical variable, using two time-point-level indicators to indicate whether the time point was at medium- or long-term, respectively. A short-term time point then corresponds to setting both these indicators to zero. We therefore assume that the effect of follow-up time is common across trials and interventions. For studies that reported outcomes at more than one follow-up time within a particular category (short-, medium- or long-term follow up), we selected the observation closest to the mid-point of the short- and medium-term intervals, and closest to 24 months for long-term observations.

In our Cochrane reviews [6, 7], we conducted risk-of-bias (RoB) assessments for each trial result using the RoB 2 tool. Here, we coded these assessments, defining a dichotomous time-point-level indicator for whether each was judged to be at high risk of bias. Separate assessments were available for each time point, so the RoB indicator depends on both the trial and the follow-up time.

### 2.5 Data analysis

#### 2.5.1 Mapping of outcome to common measurement scale

We chose zBMI as our primary outcome because, unlike BMI, it accounts for the age and sex of the child and, unlike percentile, it was reported in a large number of trials. For trials that only reported results in terms of unstandardized BMI or BMI percentiles, we mapped these values onto the zBMI scale using methods we developed previously [9]. To map from BMI to zBMI we used a sampling method, implemented with 10,000 samples. The relationship between BMI and zBMI depends on an individual’s age and sex. Assuming a lognormal distribution for BMI, a normal distribution for age and a binomial distribution for sex, we set parameters of these distributions according to information reported from the trial. We then sampled 10,000 individuals from these distributions, calculated zBMI and used these to determine mean zBMI. To obtain zBMI from BMI percentile we employed an analytic method that, assuming a normal distribution for zBMI, uses standard integral results to evaluate the expectation and variance of BMI percentile. For details of these mapping methods we refer readers to our paper [9]. In addition, nine trials reported results as the proportion of overweight or obese. As described in our Cochrane reviews, we used normality assumptions to estimate mean zBMI from these values [6, 7].

As a sensitivity analysis, we fitted the model for (non-mapped) BMI and zBMI observations separately (but not for BMI percentile due to the small number of data points). Because the conversion from BMI percentile to zBMI involves fewer assumptions than the conversion from BMI, we also performed our analysis with zBMI and (mapped) BMI percentile measurements only. In our analysis protocol we specified that we would include an indicator for whether the outcome had been mapped. Since the purpose of this is fulfilled by the sensitivity analyses, and to keep the number of variables to a minimum, we chose not to include this indicator in our model.

#### 2.5.2 Statistical model

To analyse our data we used a bespoke multi-level meta-regression model described in detail previously [10]. The model includes indicator variables (as covariates) defined on three levels: trial, intervention arm and time point. It assumes additive effects of all indicators while allowing for interactions between or within any level. An intercept term is included to capture the effect, relative to control, of an intervention whose indicators are all equal to zero. The mathematical details of our model are provided in Section B of the Supplementary Material.

Each observation in the data is a mean difference between an intervention and a reference arm. The model takes a different form depending on whether the reference arm is a control arm (control comparison) or another active intervention (active comparison). Assuming transitivity between intervention effects, the model for active comparison trials is constructed by taking the difference between two control comparison models in the same trial and at the same time point but with different intervention arms. Terms that do not depend on the intervention cancel out in this subtraction, meaning the active comparison model does not include an intercept, trial-level indicators or time-point-level indicators. Based on this construction, all regression coefficients are defined with reference to a control arm.

To account for correlations due to multi-arm and multi-follow-up trials, we specify a within-trial covariance matrix that depends on the correlation coefficient between observations at different time points. Based on observations in the data, we chose a correlation of 0.8 for the main analysis and performed sensitivity analyses with correlations of 0.5 and 0.95. For further details, refer to the Supplementary Material (Section C).

In our primary model we included random effects (RE) to capture variation in intervention effects between trials. We assumed equal between-trial heterogeneity variances across interventions and follow-up times. That is, we made the usual assumption from network meta-analysis that the variation in relative intervention effects between trials is the same for different intervention comparisons [12]. We made the additional assumption that the variation in intervention effects between trials is the same at different follow-up times, essentially treating multi-follow-up trials in the same way as multi-arm trials. As a secondary analysis, we performed a fixed effects (FE) model by setting the heterogeneity to zero. Although this was not pre-specified in our analysis plan, fixed-effects meta-regression analyses have a valid interpretation in the presence of unexplained heterogeneity [13].

We fitted our models in a Bayesian framework using Markov chain Monte Carlo (MCMC) methods implemented in JAGS [14]. Unless otherwise stated, we assigned uninformative prior distributions to all parameters. For the heterogeneity parameter, this deviated from our protocol in which we proposed to use an informative prior. However, we were able to include more data than expected in our final analysis and had sufficient information to estimate heterogeneity from the data. To aid convergence we centred all indicators (including interactions) about their mean. While this does not affect the interpretation of any estimated coefficients, it means the intercept represents the effect of indicators at their mean value. Details of the implementation of our analyses can be found in Section D of the Supplement.

#### 2.5.3 Selection of characteristics and interactions

To ensure maximum statistical power, whilst retaining the detail of our analytic framework, we aimed to reduce the number of indicators in our model through a three-step selection process. The first two steps involved the selection of indicators, while the latter focused on selecting interactions. First, we assessed the collinearity between the different indicators and inspected pairs with absolute correlations of at least 0.5. We either discarded one of the indictors or combined them into a new indicator encapsulating information from both. In a second step, we identified indicators that received identical responses (ones or zeros) more than 80% of the time. We discarded or redefined these indicators, ensuring that we retained any deemed to be particularly important by our stakeholders (see Section 2.7).

To select interaction terms, we used a Bayesian stochastic search variable selection (SSVS) approach based on work by Efthimiou et al [15]. At each iteration of the MCMC, the SSVS model uses information from the data to select interactions from a prespecified set. Essentially, the model ‘shrinks’ parameter estimates in order to avoid overfitting. Interactions for which there is the most evidence of an effect are selected most often. Given the number of indicators in our model, there were almost 300 possible pairwise interaction terms. As it was infeasible to investigate all these interactions, even within the SSVS framework, we instead focused on interactions with age and behaviour targeted (diet and/or physical activity) as these were the interactions deemed most interesting by our stakeholders (see below). We applied the SSVS model in a stepwise process, fitting models with the following interaction terms: (i) no interactions, (ii) interactions between age and all other indicators, (iii) interactions with the behaviour targeted and all other indicators. In our final model we included interactions from (ii) and (iii) that were selected more than 50% of the time. We describe details of the SSVS models in Section E of the Supplementary Material.

In presenting the results, we provide the estimated coefficient and 95% credible interval associated with each indicator and interaction term, along with the probability that each coefficient is less than zero, P(< 0), and the probability that it is greater than zero, P(>0). These probabilities were calculated as the proportion of parameter samples falling either side of zero during the MCMC (after convergence of the chain).

### 2.6 Combinations of indicator values

We evaluated the predicted outcome for every possible plausible combination of values for the indicators selected for inclusion in the model. We excluded unrealistic combinations such as a time point being both medium and long term, or an intervention targeting ‘physical activity alone’ and ‘both diet and physical activity’. We also ensured that at least one intervention-level indicator for a mechanism of action (participation, education, social environment or physical environment) was non-zero. The outcome of the model is in units of mean difference in change from baseline in zBMI (intervention relative to control). Therefore, the smaller (more negative) the model outcome, the more beneficial the intervention. To identify which combination of indicator values predicts the best results (greatest effectiveness), we searched for the instance associated with the minimum value of the outcome. To interpret the magnitude of effect at this combination, we converted the intercept to its equivalent value for non-centred indicators using the mean value of each indicator (see Section F of the Supplement for details).

### 2.7 Patient and public involvement

We involved members of the public in multiple stages of the work. Two school attenders were members of the project advisory group. In the development of the analytic framework, we held two workshops involving 11 children and young people up to age 18 and two workshops involving eight schoolteachers. At these workshops, we generated ideas for inclusion in the analytic framework. The full analytic framework was later discussed in a larger meeting including one young person and one schoolteacher from these workshops. We additionally involved 35 children and young people (ages 6 to 18) in the coding of the interventions; relying on them for all coding decisions of the ‘fun factor’ item [8].

## 3 Results

Among the 244 trials included in the Cochrane reviews, only 208 provided data that allowed their inclusion in meta-analyses and four of these provided data in a form that could not be converted to zBMI. Our analysis included 295 observations from 255 intervention arms in the remaining 204 trials. The trials either reported zBMI data directly (171 observations from 110 trials) or provided data that could be mapped to zBMI from BMI (88 observations from 67 trials), percentiles (25 observations from 18 trials) or proportions of the intervention arm in different weight categories (11 observations from 9 trials). The observations correspond to 250 comparisons between an intervention and a reference arm, some of which were observed at multiple time points. Brief characteristics of the trials included in our analysis are summarized in Table S5 in Section G of the Supplementary Material; further details can be found in our two Cochrane reviews [6, 7].

### 3.1 Selection of indicator variables

The final indicators included in the model are listed in Table 1 with a brief description of how they were coded and the results of the coding. We included 18 intervention-level, three trial-level and three time-point-level indicators. The coded characteristic ‘change of behaviour targeted’ encapsulated whether the intervention targets diet alone, physical activity alone or a combination of both; we coded this as two indicators, treating ‘diet alone’ as a reference. In Section H of the Supplementary Material, we describe the results of our indicator selection process, including the correlation matrix between the intervention-level indicators, the proportion of indicators with identical responses, and which indicators were removed or redefined at each step.

**Table 1.** Description of indicators included in the final analysis.

| Indicator variable | Description | Coding | n (%) |
| --- | --- | --- | --- |
| **Intervention-level indicators (n=250 included arms)** |  |  |  |
| School | Was the intervention delivered in a school (in full or in part)? | 1 if Yes<br>0 if No | 177 (70.8%)<br>73 (29.2%) |
| Home | Was the intervention delivered in the home (in full or in part) OR did it include any home activity for the child? | 1 if Yes<br>0 if No | 106 (42.4%)<br>144 (57.6%) |
| Community | Was the intervention delivered in the community or other setting (in full or in part)? | 1 if Yes<br>0 if No | 70 (28.0%)<br>180 (72.0%) |
| Individual | Was the intervention delivered to the child individually (either exclusively OR both individually and as part of a group)? | 1 if Yes<br>0 if No | 119 (47.6%)<br>131 (52.4%) |
| Electronic | Did the intervention involve any electronic component (exclusively/significantly/as a minor component)? | 1 if Yes<br>0 if No | 53 (21.2%)<br>197 (78.8%) |
| Diet and physical activity | Did the intervention aim to change both diet and activity? | 1 if Yes<br>0 if No | 137 (54.8%)<br>113 (45.2%) |
| Physical activity | Did the intervention aim to change physical activity alone? | 1 if Yes<br>0 if No | 68 (27.2%)<br>182 (72.8%) |
| Multi-strategy | Did the intervention use multiple strategies (three or more)? | 1 if Yes<br>0 if No | 161 (64.4%)<br>89 (35.6%) |
| Duration | Was the intervention long ($\geq 30.33$ weeks) or short ($< 30.33$ weeks)?* | 1 if Long<br>0 if Short | 125 (50.0%)<br>125 (50.0%) |
| Intensity | What was the level of engagement with the children during the peak engagement period?*** | 1 if High<br>0 if Low | 152 (60.8%)<br>98 (39.2%) |
| Integration | Was the intervention integrated into the normal curriculum/ habits? | 1 if Yes<br>0 if No/<br>Partially | 118 (47.2%)<br>132 (52.8%) |
| Flexibility/choice | Was the intervention designed to be implemented in a flexible manner OR to include choice for the child? | 1 if Yes<br>0 if No | 116 (46.4%)<br>134 (53.6%) |
| Fun factor | Was the intervention considered fun? | 1 if Fun<br>0 if Boring/<br>Neutral | 153 (61.2%)<br>97 (38.8%) |
| Resonance | Was the intervention experienced by children via someone external or unusual? | 1 if Yes<br>0 if No | 131 (52.4%)<br>119 (47.6%) |
| Participation | Did the intervention have an explicit component of modifying the child's behaviour? | 1 if Yes<br>0 if No | 168 (67.2%)<br>82 (32.8%) |
| Education | Did the intervention have an explicit component of education/information provision for the child? | 1 if Yes<br>0 if No | 185 (74.0%)<br>65 (26.0%) |
| Social environment | Did the intervention have an explicit component aiming to change the social environment of the child? | 1 if Yes<br>0 if No | 174 (69.6%)<br>76 (30.4%) |
| Physical environment | Did the intervention have an explicit component aiming to change the physical environment of the child? | 1 if Yes<br>0 if No | 79 (31.6%)<br>171 (68.4%) |
| **Trial-level indicators (n=204 trials)** |  |  |  |
| Age | What was the targeted age of children in the trial (based on mean age)? | 1 if 12-18 yrs<br>0 if 5-11 yrs | 54 (26.5%)<br>150 (73.5%) |
| Income country | What is the income status of country in which the trial was conducted according to World Bank criteria? | 1 if High<br>0 if Non-high | 175 (85.8%)<br>29 (14.2%) |
| SES | What was the socio-economic status of the participants (based on categorizations described by the trial authors)? | 1 if Mixed<br>0 if Low | 157 (77.0%)<br>47 (23.0%) |
| **Time-point-level indicators (n=261 observed time points)** |  |  |  |
| Medium | Was the follow-up time medium-term (9 months to <15 months)? | 1 if Yes<br>0 if No | 102 (39.1%)<br>159 (60.9%) |
| Long | Was the follow-up time long-term (>15 months)? | 1 if Yes<br>0 if No | 88 (33.7%)<br>173 (66.3%) |
| ROB | Was the result at high risk of bias? | 1 if High<br>0 if Low/Some concerns | 63 (24.1%)<br>198 (75.9%) |

\*30.33 weeks is the median duration across all interventions (including studies excluded from the primary analysis)
\*\*As described in [8], high intensity refers to the child engaging with the intervention at least once a week, low intensity reflects engagement of less than once a week.

Below, we list the interactions selected by the stepwise SSVS procedure for the primary (random effects) analysis. We underline interactions that are different between the RE and FE analyses (i.e., selected in one but not the other).

- Interactions with age: electronic, diet and physical activity, multi-strategy, integration, resonance, <u>education</u>, income status of country
- Interactions with diet and physical activity: electronic, <u>multi-strategy</u>, fun factor, <u>resonance</u>, age [as above], <u>risk of bias</u>
- Interactions with physical activity only: electronic, duration, <u>fun factor</u>, income status of country, risk of bias

Interactions selected for the secondary (fixed effects) analysis were:

- Interactions with age: electronic, diet and physical activity, multi-strategy, integration, <u>fun factor</u>, resonance, income status of country
- Interactions with diet and physical activity: <u>school, home</u>, electronic, fun factor, <u>social</u>, age [as above], <u>income status of country</u>
- Interactions with physical activity only: <u>community, individual</u>, electronic, <u>multi-strategy</u>, duration, income status of country, risk of bias

For the results of the SSVS model at each step, see Section I of the Supplementary Material.

### 3.2 Main results

#### 3.2.1 Primary analysis (random effects)

Figure 1 shows a forest plot of the results of our primary analysis (random effects model). Intervention effects were measured as mean differences (MD) in change from baseline in zBMI. Since effective interventions lead to smaller increases (or larger decreases) in zBMI, coefficients of indicators that are less than zero indicate greater effectiveness. The estimate of the model intercept and its 95% credible interval was −0.037[−0.053, −0.022]. This represents the effect of an intervention with all indicators set to their mean value, indicating that the interventions were beneficial on average. The heterogeneity standard deviation (and its 95% credible interval) was estimated to be 0.080 [0.067, 0.093], which is relatively large compared with the typical values of the estimated coefficients. In the following we present estimates of coefficients as differences in mean differences and their 95% credible intervals (DMD [95% CI]). For a particular indicator, the DMD represents the additional effect of an intervention with indictor value 1 compared with an intervention with indicator value 0, conditional on the two interventions sharing the same values of all other indicators.

**Figure 1.**
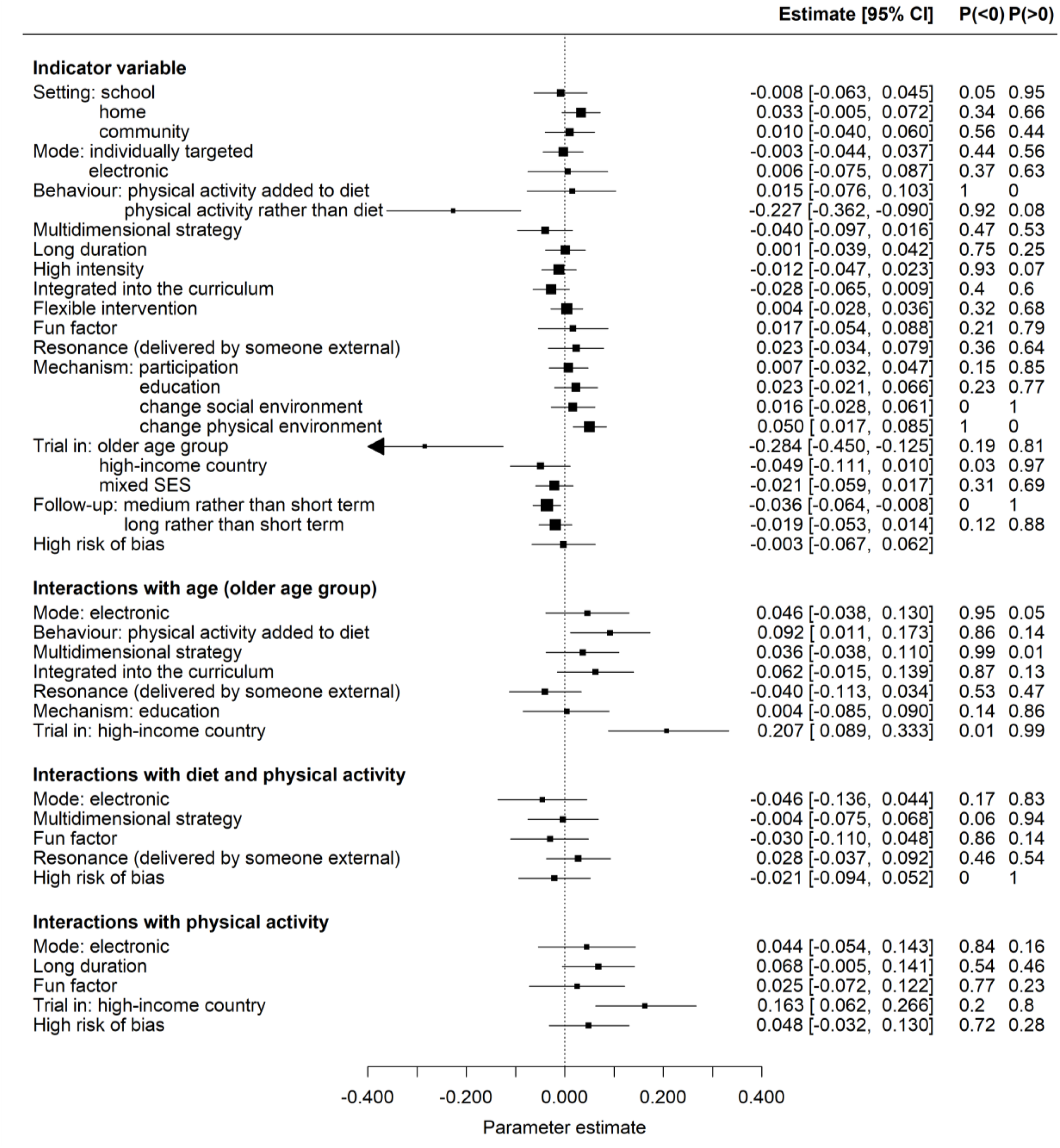
Parameter estimates from the primary (random effects) analysis. On the right, we list the estimate of each regression coefficient and its 95% credible interval along with the probability that the coefficient is less than or greater than zero, *P*(< 0) and *P*(>0). Intervention effects were measured as mean differences (MD) in change from baseline in zBMI therefore coefficients of indicators that are less than zero indicate greater effectiveness.

Our results provide strong evidence for a greater beneficial effect of interventions targeting physical activity alone compared with diet alone (−0.227 [−0.362, −0.090], P(< 0) = 1). Conversely, we observe no evidence of differing effects between diet and physical activity compared with diet alone. There is some indication of (small) greater effects for interventions that use multiple strategies (−0.040 [−0.097, 0.016], P(< 0) = 0.92) and are fully integrated within the curriculum/every day habits (−0.028 [−0.065, 0.009], P(< 0) = 0.93). On the other hand, the results indicate that interventions are less beneficial if they involve a change to the physical environment (most commonly the food environment) (0.050 [0.017, 0.085], P(>0) = 1) and possibly less beneficial if they have some home-based component (0.033 [−0.005, 0.072], P(>0) = 0.95).

There is strong evidence that, overall, interventions are more effective for children aged 12-18 years than for those aged 5-11 years (−0.284 [−0.450, −0.125], P(< 0) = 1). The analysis also suggests that the average intervention may work better in higher income countries (−0.049 [−0.111, 0.010], P(< 0) = 0.95). However, both age and income status of country appear in multiple interactions so the interpretation of these results is more complicated. We return to this in Section 3.2.3. The results for follow-up time indicate that, compared with short-term, larger beneficial effects are seen at medium-term (−0.036 [−0.064, −0.008], P(< 0) = 0.99), potentially followed by long-term (−0.019 [−0.53, 0.014], P(< 0) = 0.87).

Interactions are described as synergistic if the combined effect of the two indicators is greater than the sum of each independently or as antagonistic if the combined effect is less than the sum of each independently. In Figure 1, interactions less than zero indicate a synergistic effect of the two indicators whereas interactions greater than zero indicate an antagonistic effect. We find antagonistic interactions between the older age group and (i) interventions targeting both diet and physical activity, and (ii) interventions conducted in high income countries. We also observe antagonistic interactions between interventions targeting physical activity alone and (i) those conducted in high income countries, and (ii) interventions with long duration. We find no evidence of interactions with interventions that target both diet and physical activity. We discuss the interpretation of these interactions in Section 3.2.3.

#### 3.2.2 Secondary analysis (fixed effects)

The results of our secondary analysis (fixed effects model) are shown in Figure 2. As expected, this analysis leads to more precise parameter estimates. In addition to the effects identified in the primary analysis, the FE model suggests beneficial effects for interventions that are based in a school, target diet and physical activity (compared with diet alone) and are high intensity. It also identifies additional, less beneficial, effects for interventions that are considered fun, resonant and involve an educational component. The FE analysis provides stronger evidence (compared with RE) that the average effect of interventions is more beneficial in higher income countries, and that greater benefits are seen at long term follow-up compared to short term. In addition, this model finds that high risk of bias is associated with larger beneficial effects.

**Figure 2.**
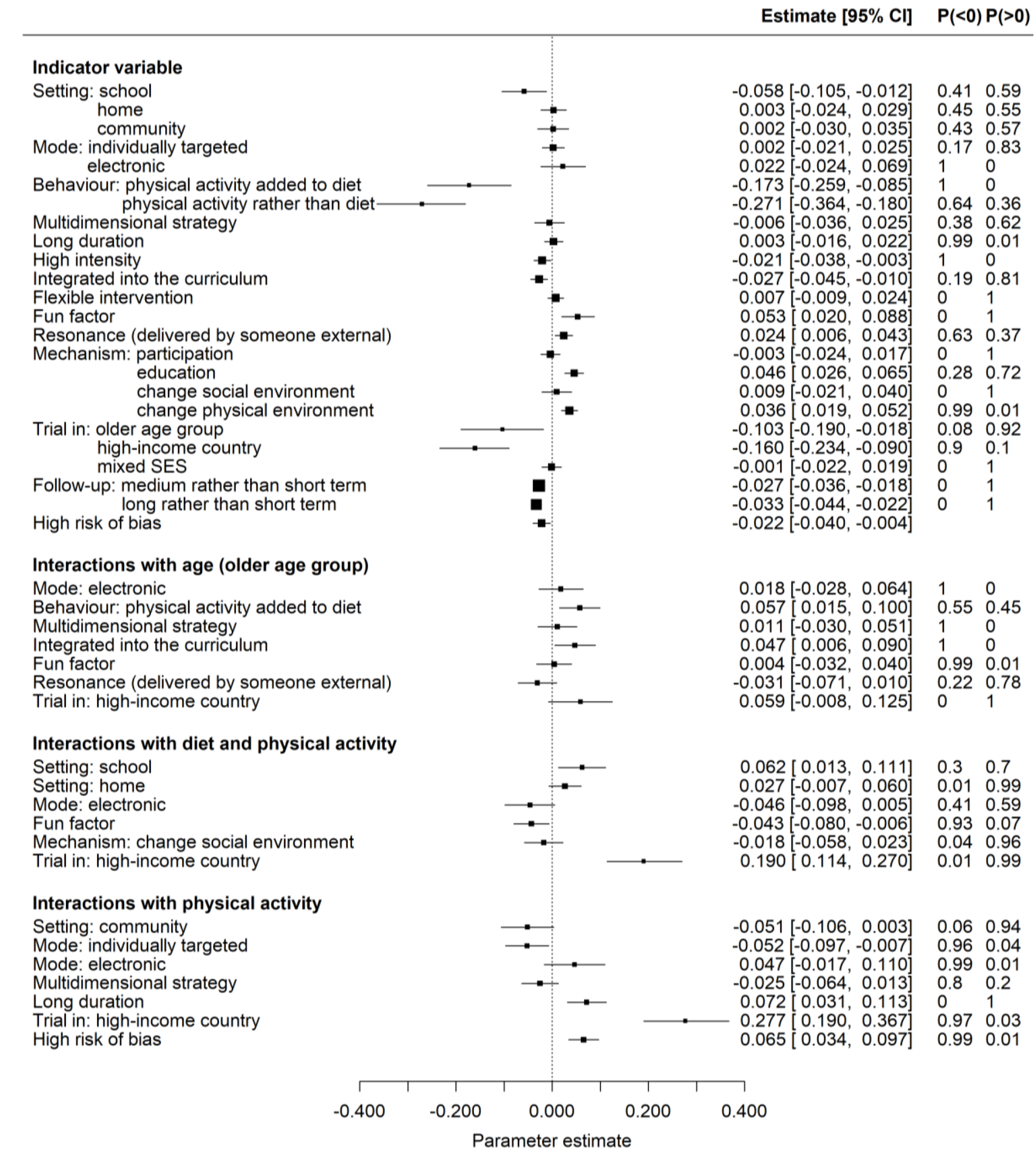
Parameter estimates from the secondary (fixed effects) analysis. On the right, we list the estimate of each regression coefficient and its 95% credible interval along with the probability that the coefficient is less than or greater than zero, *P*(< 0) and *P*(>0). Intervention effects were measured as mean differences (MD) in change from baseline in zBMI therefore coefficients of indicators that are less than zero indicate greater effectiveness.

In addition to the interactions identified by the RE analysis, the FE model identifies several other effects. For the older age group, we observe antagonistic interactions with interventions that are integrated into the curriculum, and, with lower certainty, synergistic effects with interventions delivered by someone resonant. For interventions targeting both diet and physical activity, the FE model finds strong evidence of antagonistic effects with school-based interventions and high-income countries, and weaker evidence for antagonistic effects with interventions than involve home-based activities or delivery. Conversely, we observe synergistic effects between diet and physical activity interventions and those that are considered fun and those that involve an electronic component. For interventions that target physical activity alone, we find reasonable evidence of all interactions (probabilities ≥ 0.9). The strongest signals are for antagonistic effects with long duration, high income countries, and high risk of bias, and synergistic effects with interventions delivered in a community setting and those involving an individual-level component.

#### 3.2.3 Combinations of indicators

By evaluating the model at every combination of indicator values, we found that it predicts the greatest effectiveness for interventions that are: set in a school, have an element that is delivered individually, target physical activity alone, contain multiple strategies, are high intensity, of short duration, are delivered by someone resonant, and aim to modify behaviour through participation. These findings are associated with the older age group, low-income countries, mixed (rather than low) socio-economic status and medium-term follow-up. Since we found evidence of various interactions with age and income status, we re-evaluated the ‘best’ combination of indicator values specific to each age group in both high and non-high-income countries.

Table 2 summarizes these results. For the younger age group, resonance is not associated with the largest beneficial effect, but instead the model identifies interventions that are integrated into the curriculum or daily habits. The indicator combination which differs most from all others is for the younger age group in high income countries. This is the subset associated with the smallest most beneficial effect. In contrast to the older age group and the younger age group in low-income countries, the results indicate that beneficial effects in this group are associated with interventions involving an electronic component, that target both diet and physical activity and are fun.

**Table 2.**
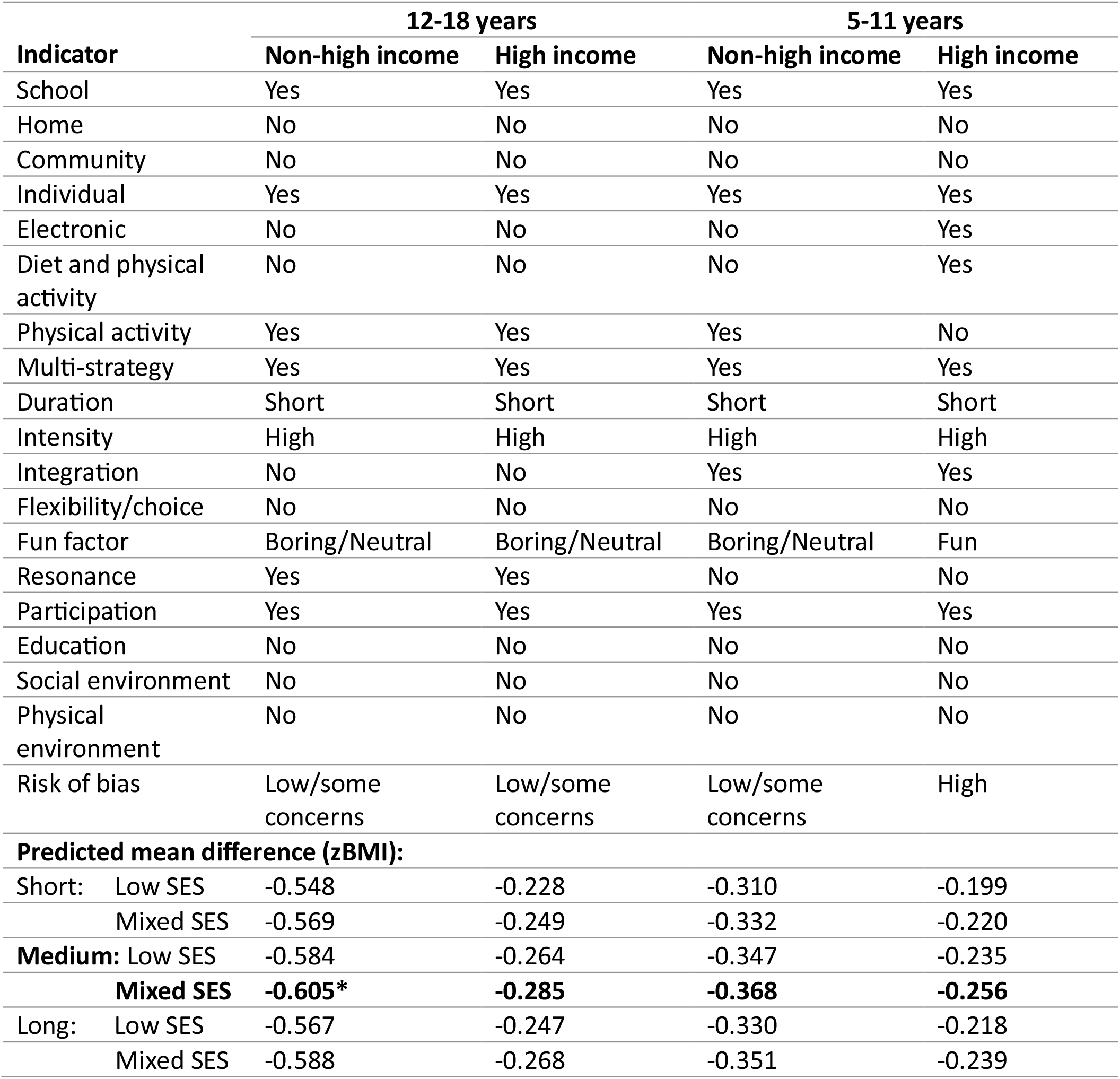
The indicator values associated with the minimum (most negative) mean difference predicted from our primary (random-effects) model. We show the most beneficial combination of indicator values for each of the two age groups (5-11 years and 12-18 years) and the two levels of country income status (high and non-high). We find the same combination regardless of whether we specify follow-up time (short, medium or long) or SES (mixed or low). The resulting predicted mean difference in each scenario is shown at the bottom of the table. For a given age group and income status, the largest beneficial effect is obtained for medium term follow-up and mixed SES (or universal interventions; highlighted in bold). Overall, the model predicts the greatest benefit for the older age group in non-high-income countries (indicated with an asterisk).

| Indicator | 12-18 years |  | 5-11 years |  |
| --- | --- | --- | --- | --- |
|  | Non-high income | High income | Non-high income | High income |
| School | Yes | Yes | Yes | Yes |
| Home | No | No | No | No |
| Community | No | No | No | No |
| Individual | Yes | Yes | Yes | Yes |
| Electronic | No | No | No | Yes |
| Diet and physical activity | No | No | No | Yes |
| Physical activity | Yes | Yes | Yes | No |
| Multi-strategy | Yes | Yes | Yes | Yes |
| Duration | Short | Short | Short | Short |
| Intensity | High | High | High | High |
| Integration | No | No | Yes | Yes |
| Flexibility/choice | No | No | No | No |
| Fun factor | Boring/Neutral | Boring/Neutral | Boring/Neutral | Fun |
| Resonance | Yes | Yes | No | No |
| Participation | Yes | Yes | Yes | Yes |
| Education | No | No | No | No |
| Social environment | No | No | No | No |
| Physical environment | No | No | No | No |
| Risk of bias | Low/some concerns | Low/some concerns | Low/some concerns | High |
| **Predicted mean difference (zBMI):** |  |  |  |  |
| Short: Low SES | -0.548 | -0.228 | -0.310 | -0.199 |
| Mixed SES | -0.569 | -0.249 | -0.332 | -0.220 |
| Medium: Low SES | -0.584 | -0.264 | -0.347 | -0.235 |
| **Mixed SES** | **-0.605\*** | **-0.285** | **-0.368** | **-0.256** |
| Long: Low SES | -0.567 | -0.247 | -0.330 | -0.218 |
| Mixed SES | -0.588 | -0.268 | -0.351 | -0.239 |

For each subset (age and income status) the largest beneficial effect predicted by the model is for medium-term follow-up and mixed socio-economic status (rather than targeted at low socio-economic status). We re-evaluated the most beneficial indicator combination specific to low socio-economic status, long-term follow-up and short-term follow-up respectively. In each subset, the model identified the same set of intervention indicators associated with the largest beneficial effect. In Table 2 we list the value of the model obtained in each scenario.

#### 3.2.4 Sensitivity analyses

Sensitivity analyses with different outcome scales analysed separately, i.e., excluding any mapped data, are reported in full in Section J of the Supplementary Material. Results for BMI only are similar to those of the primary analysis, while the zBMI only results are largely uncertain, with wider credible intervals. The only conflicting evidence we observe is for the average effect of high-income countries: for BMI alone effects are greater in high-income countries, while for zBMI alone there is a greater beneficial effect in low-income countries; this could be due in part to the small amount of data available from low-income countries. For the interaction terms, we observe no conflicting evidence between the analyses on separate outcome scales. The only interaction that appears with strong evidence in one of the separate outcome analyses that does not appear in the primary analysis is a synergistic interaction between interventions with an electronic component and interventions targeting both diet and physical activity in the analysis of BMI only. In sensitivity analyses of the main random effects analysis assuming different correlations between observations at different time points we observed negligible differences from the primary analysis across all estimated effects (Section K of the Supplementary Material).

## 4 Discussion

In our re-analysis of the results of 204 randomized trials using an analytic framework co-developed with stakeholders, we found that the most effective characteristic to include in a behaviour-change intervention to prevent obesity in children was physical activity. Physical activity interventions delivered in a school setting, that included active participation, were of high intensity and short duration and were delivered through multiple strategies appeared the most effective. For young children (of primary school years) living in high income countries, greater effectiveness appeared to be possible where these interventions were also integrated into the normal school day, included a healthy diet, involved an electronic component and were ‘fun’. Although these beneficial effects are small, when delivered at scale, the effects of these preventive interventions have the potential to contribute meaningfully to a reduction in the prevalence of childhood obesity [16].

Strengths of our investigation include our use of a large, comprehensive, updated systematic review of randomized trials; selection of indicator variables derived from a co-produced analytic framework that benefitted from the involvement of children, young people, teachers and public health professionals; careful coding of intervention and trial characteristics by a mixture of researchers and children/young people themselves; and sophisticated statistical methods. The study is not without limitations. These include our dependence on the nature of interventions that have been investigated in randomized trials, which were mainly school based and include many that would be delivered in different ways now (eligible trials were published between 1990 and 2023). For example, the role of electronic or digital implementation of interventions could not be examined in detail. Trial reports also provided very little information on how well interventions were implemented; we did not include implementation issues in our analytic framework for this reason. We did not explore in detail the impact of participant characteristics on intervention effectiveness. Average results for whole populations can hide differences in effects between subgroups of the population and these differences may lead to, or widen, health inequalities. It is important that attempts to prevent obesity in children ensure, as best they can, that they minimize inequity. In a parallel project we have examined this question in detail for the trials included here [17].

Many papers have reported the benefits of one or more of the behaviour-change characteristics considered here, although few have employed a systematic approach using controlled studies. Our main finding concurs with that of a previous study which demonstrated the effectiveness of physical activity interventions in the school setting, particularly when included in the school curriculum and emphasizing participants’ enjoyment [18]. Another study examined interventions that included diet combined with physical activity and found that effective strategies included changes in the schoolyard, in the recess rules and in the physical education classes [19].

We were surprised by our finding that modification of the physical environment was associated with an unfavourable impact on prevention strategies, given the general understanding that this should be useful. Most of the modifications used in the interventions in our study related to the food environment, either alone or alongside changes to the physical activity environment. This is however consistent with findings of a previous study which found only two (of nine) studies employing interventions aiming to modify the food and built environments within and around schools were effective [20]. We also observed, unexpectedly, a suggestion that the inclusion of a home activity is not useful [21]. However, we did not assess the degree of active parental involvement and in most of the trials this only extended to newsletters and other educational information sent to the home of the child.

Much effort is invested by governments globally in childhood obesity prevention policies that address food and beverages. For example, in England, current headline actions include a soft drinks industry levy (‘sugar tax’), calorie labelling, town planning restrictions for hot food take-aways and partial banning of advertisements for less healthy products on television, with much less focus on the promotion of physical activity (a notable exception being funding for schools to support efforts in promoting physical education and engagement with sport). We suggest that even greater gains might be achievable if actions were also focussed on promoting physical activity. In a similar vein, most schools have either separate policies (or programmes) around food and physical activity/sport or an overarching policy for both, and these have been found to afford relatively more attention to food compared with physical activity [22]. Our findings are particularly relevant to those providing guidance for schools and we encourage those responsible to ensure strategies relating to physical activity are as comprehensive as those for food.

There is increasing enthusiasm for applying ‘whole-systems’ approaches to communities, societies and schools to address childhood obesity [23]. These highlight the importance of upstream interventions and those requiring lower individual agency as the key to success. A whole-systems approach involves multiple strategies and levels of intervention interconnected via a programme theory and logic model. Included in these are specific strategies, often school based, that are of the type included in the two Cochrane reviews feeding into this work. We believe the findings of this work are therefore relevant to those providing guidance on, or implementing, whole-systems approaches to preventing obesity in children and young people.

Being physically active and consuming a healthy diet during childhood offer many important benefits beyond contributing to healthy weight and development, including well-being and mental health, dental health, the ability to learn and educational attainment, and realization of full life-time potential [4]. The findings presented in this paper should not be misinterpreted as ‘diet doesn’t matter’; it does. Our findings suggest that behaviour change interventions to prevent obesity in children should focus primarily on the promotion of physical activity and should consider the other effective characteristics we identify here.

## Data Availability

All data and codes that support the findings of this study are available at the GitHub repository: https://github.com/AnnieDavies/Obesity_Synthesis

## Ethics statement

Ethical approval: none required.

## Acknowledgements

ALD would like to thank Hugo Pedder for useful discussions related to the implementation of the statistical analysis.

The authors would like to thank our PPI contributors, specifically Maddy Coleman and Elizabeth Sheldrick for their role as the children and young people representatives in the project advisory group.

## Author contributions

Annabel L Davies: writing – original draft, formal analysis (lead), methodology (equal), data curation (equal), software (lead), visualization (lead), writing – review & editing (lead), investigation (data acquisition and coding) (equal)

Francesca Spiga: data curation (equal), investigation (study identification, data acquisition and coding) (lead), writing – review & editing (supporting)

Deborah M Caldwell: conceptualization (equal), funding acquisition (supporting), methodology (equal), investigation (data acquisition and coding) (equal), writing – review & editing (supporting)

Jelena Savović: funding acquisition (supporting), investigation (data acquisition and coding) (equal), writing – review & editing (supporting)

Jennifer Palmer: investigation (study identification, data acquisition and coding) (equal), writing – review & editing (supporting)

Eve Tomlinson: investigation (study identification, data acquisition and coding) (equal), writing – review & editing (supporting)

Theresa HM Moore: investigation (study identification, data acquisition and coding) (equal), writing – review & editing (supporting)

Carolyn D Summerbell: funding acquisition (lead), supervision (equal), writing – review & editing (lead), investigation (study identification, data acquisition and coding) (equal)

Julian PT Higgins: conceptualization (equal), methodology (equal), project administration (lead), supervision (equal), funding acquisition (lead), writing – review & editing (lead), investigation (data acquisition and coding) (equal)

The corresponding author attests that all listed authors meet authorship criteria and that no others meeting the criteria have been omitted.

## Transparency

Dr Davies affirms that this manuscript is an honest, accurate, and transparent account of the study being reported; that no important aspects of the study have been omitted; and that any discrepancies from the study as originally planned have been explained.

## Dissemination to participants and related patient and public communities

The results of the study were discussed with public contributors at two meetings dedicated to this in January 2024 to help us disseminate the findings and interpret the results. The first meeting involved five children and young people. The second meeting involved four current and former teachers. In addition, one public contributor from each of these workshops participated in an expert meeting with public health professionals in February 2024 where we presented the results and invited comment.

## Supplementary material

### A Coding of binary intervention indicators

Below we list the (dichotomous) intervention indicators defined by the analytic framework and how they were coded. For details of how the questions were answered, we refer to [1]. We describe any dichotomizations applied to continuous or categorical variables and give the resulting split of the data (where we write *n* for the number of interventions that fall into that category). The numbers refer to the full set of coded interventions (*n* = 255) as reported in [1] rather than the final set of interventions included in the analysis (*n* = 250, see Table 1 in the main paper).

1. School: was the intervention delivered in a school (in full or in part)?
  ∘ = 1 if Yes (*n* = 180)
  ∘ = 0 if No (*n* = 75)
2. Home: was the intervention delivered in the home (in full or in part)?
  ∘ = 1 if Yes (*n* = 47)
  ∘ = 0 if No (*n* = 208)
3. Community: was the intervention delivered in the community or other non-school/non-home setting (in full or in part)?
  ∘ = 1 if Yes (*n* = 72)
  ∘ = 0 if No (*n* = 183)
4. Home activity: did the intervention include a home activity for the child?
  ∘ = 1 if Yes (*n* = 91)
  ∘ = 0 if No (*n* = 164)
5. Group: Was the intervention delivered to the child as part of a group (in full or in part)?
  ∘ = 1 if Yes (*n* = 211)
  ∘ = 0 if No (*n* = 44)
6. Individual: Was the intervention delivered to the child individually (in full or in part)?
  ∘ = 1 if Yes (*n* = 122)
  ∘ = 0 if No (*n* = 133)
7. Electronic: was the intervention delivered to the child electronically?
  ∘ = 1 if Yes exclusively/Yes significantly/Yes as a minor component (*n* = 54)
  ∘ = 0 if No (*n* = 201)
8. Diet: Did the intervention aim to change diet?
  ∘ = 1 if Yes exclusively/Yes substantially (*n* = 187)
  ∘ = 0 if No/Yes minimally (*n* = 68)
9. Physical activity: Did the intervention aim to change physical activity?
  ∘ = 1 if Yes exclusively/Yes substantially (*n* = 207)
  ∘ = 0 if No/Yes minimally (*n* = 48)
10. Multi-strategy: Did the intervention use multiple strategies (three or more)?
  ∘ = 1 if Yes (*n* = 161)
  ∘ = 0 if No (*n* = 94)
11. Single phase: was the intervention applied in a single phase?
  ∘ = 1 if Yes (*n* = 207)
  ∘ = 0 if No (*n* = 48)
12. Continuous: was the intervention applied continuously?
  ∘ = 1 if Yes (*n* = 246)
  ∘ = 0 if No (*n* = 9)
13. Total duration: what was the total duration of the intervention?
  ∘ = 1 if Long (longer than median total duration, ≥ 30.33 weeks) (*n* = 128)
  ∘ = 0 if Short (shorter than median total duration, < 30.33 weeks) (*n* = 127)
14. Peak duration: what was the duration of the period of peak engagement with the intervention?
  ∘ = 1 if Long (longer than median peak duration, ≥ 25.98 weeks) (*n* = 131)
  ∘ = 0 if Short (shorter than median peak duration, < 25.98 weeks) (*n* = 124)
15. Intensity: what was the level of engagement with the children during the peak period?
  ∘ = 1 if High (*n* = 152)
  ∘ = 0 if Low (*n* = 103)
16. Integration: was the intervention integrated into the normal curriculum/ habits?
  ∘ = 1 if Yes (*n* = 121)
  ∘ = 0 if No/Partially (*n* = 134)
17. Flexibility: was the intervention designed to be implemented in a flexible manner/tailored to specific participants?
  ∘ = 1 if Yes (*n* = 86)
  ∘ = 0 if No (*n* = 169)
18. Choice: was choice for the child of physical activity/diet designed into the intervention?
  ∘ = 1 if Yes (*n* = 66)
  ∘ = 0 if No (*n* = 189)
19. Fun factor: was the intervention considered fun?
  ∘ = 1 if Fun (*n* = 154)
  ∘ = 0 if Neutral/Not fun (*n* = 101)
20. Resonance: was the intervention experienced by children via someone external or unusual?
  ∘ = 1 if Yes (*n* = 134)
  ∘ = 0 if No (*n* = 121)
21. Participation: did the intervention have an explicit component that requires the child to participate?
  ∘ = 1 if Yes (*n* = 170)
  ∘ = 0 if No (*n* = 85)
22. Education: did the intervention have an explicit component of education/information provision for the child?
  ∘ = 1 if Yes (*n* = 190)
  ∘ = 0 if No (*n* = 65)
23. Social: did the intervention have an explicit component aiming to change the social environment of the child (e.g., at school or home)?
  ∘ = 1 if Yes (*n* = 175)
  ∘ = 0 if No (*n* = 80)
24. Environment: did the intervention have an explicit component aiming to change the physical environment of the child (e.g., at school or home)?
  ∘ = 1 if Yes (*n* = 79)
  ∘ = 0 if No (*n* = 176)
25. Commercial: were commercial interests involved in the intervention/trial?
  ∘ = 1 if Yes (*n* = 27)
  ∘ = 0 if No (*n* = 228)

### B Mathematical details of the complex synthesis model

#### B.1 Random effects model

Each trial *i* with *A*_*i*_ arms and *T*_*i*_ follow-up times provides a *T*_*i*_(*A*_*i*_ − 1) × 1 vector of observations 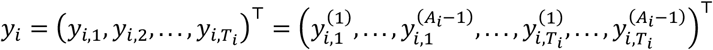 . Each 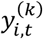 is a mean difference in change from baseline to time *t* between arm *k* and some trial-specific reference arm *r*. The reference arm is either a control arm, *r* = *C*, or another active intervention, *r* = *A*_*i*_. As described in our Cochrane reviews [2, 3], we calculated mean differences from arm-level data selected in the following order of preference: (i) follow-up means adjusted for baseline values, (ii) mean change from baseline (change scores), (iii) unadjusted baseline and follow-up means, (iv) unadjusted follow-up means without baseline data. For scenario (iii) we used the baseline and follow-up values to first calculate change from baseline. To obtain standard errors on these change scores we assumed a correlation coefficient of 0.9 between baseline and follow-up based on observed correlations in other trials. This is different from the value of 0.95 we used in our Cochrane reviews which was informed by initial observations of a subset of trials.

We define a meta-regression style model assuming additive effects of indicator variables (covariates) on three levels: study (*z*_*i*_), intervention 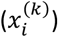, and follow-up time (*w*_*i*,*t*_). We also include pairwise interaction terms 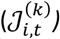 between indicators on any level. The form of the model depends on the reference arm of the trial and is summarized as follows,

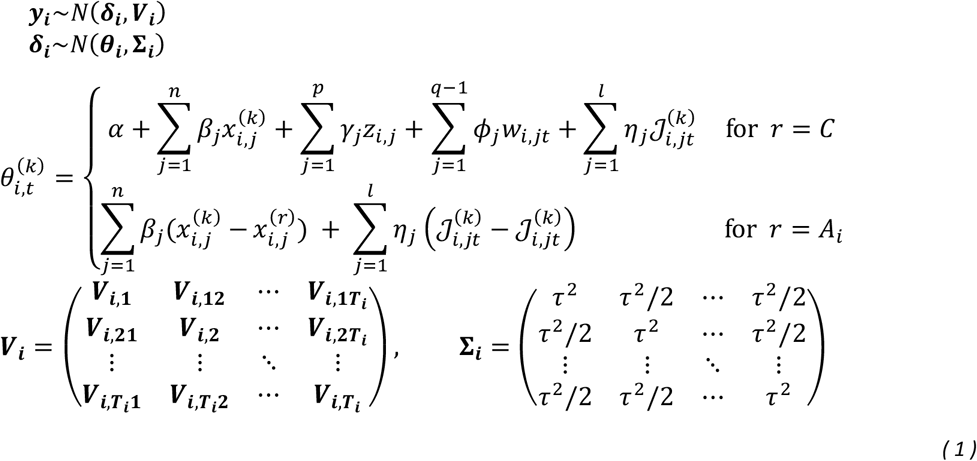

where the parameters *β* = (*β*_1_ , … , *β*_*n*_)^⊤^, *γ* = (*γ*_1_ , … , *γ*_*p*_ )^⊤^, and *ϕ* = (*ϕ*_1_ , … , *ϕ*_*q*−1_)^⊤^are the regression coefficients for the intervention, study-level, and follow-up time indicators respectively. The parameters *η* = (*η*_1_, … , *η*_*l*_)^⊤^ are the regression coefficients for the interaction parameters.

The *T*_*i*_(*A*_*i*_ − 1) × *T*_*i*_(*A*_*i*_ − 1) covariance matrix ***V***_***i***_ captures correlations between measurements made in the same trial (due to multiple arms and/or follow-up times) and is assumed to be known. The diagonal elements of the covariance matrix are equal to the observed variance (squared standard error) of the measurements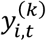. The off-diagonal elements capture correlations between multiple observations in the same trial and depend on two (imputed) correlation coefficients *ρ*_*y*,*tt*′_ and *ρ*_*d*,*tt*′_ (described in Section C). We refer to our development paper [4] for details of this matrix.

For the random effects model, we assume the trial-specific effects 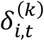 are normally distributed with covariance matrix Σ_***i***_ (dimensions *T*_*i*_(*A*_*i*_ − 1) × *T*_*i*_(*A*_*i*_ − 1)). This matrix depends on the heterogeneity variance, *τ*^2^, which we assume captures the between-trial variance for all arms and all follow-up times.

#### B.2 Fixed effects model

As our secondary analysis we define a fixed effects model by setting *τ* = 0 in Equation ( 1 ). This yields

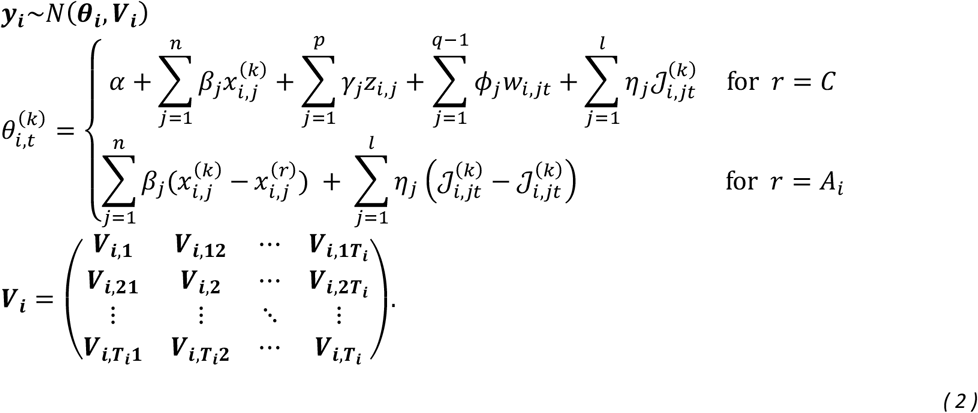

### C Imputing correlation coefficients to specify the within study covariance matrix

As described in Section B, the within-study covariance matrix in our model depends on two correlation coefficients *ρ*_*y*,*tt*′_ and *ρ*_*d*,*tt*′_. The former is the correlation between observations of mean difference for a given intervention (relative to the reference arm) at time points *t* and *t*′, assumed to be common to all arms. The latter is the correlation between observations of the change score in the reference arm between *t* and *t*′.

To specify the within-study covariance matrix, we imputed sensible values for the correlations *ρ*_*y*,*tt*′_ and *ρ*_*d*,*tt*′_ based on observed correlations in the data. Using multi-follow-up trials, we plotted mean differences and change scores in control arms for different combinations of time points: (i) short vs medium term, (ii) short vs long term, and (iii) medium vs long term. We then calculated correlation coefficients for each plot.

To simplify the imputation we assumed that time points with one degree of separation, (i) and (iii), have the same correlation, *ρ*_1_. We also assumed that correlations act multiplicatively across time such that time points with two degrees of separation, (ii), have the square of this correlation, 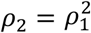.

For mean differences, *ρ*_*y*,*tt*′_, we found a correlation of *ρ*_1_ = 0.77 for short vs medium term (based on *m* = 31 data points), *ρ*_2_ = 0.83 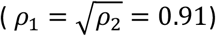 for short vs long term (*m* = 13), and *ρ*_1_ = 0.86 for medium vs long term (*m* = 45). This gives an overall weighted mean of *ρ*_1_ = 0.82.

For change scores, *ρ*_*d*,*tt*′_, we found a correlation of *ρ*_1_ = 0.69 for short vs medium term (*m* = 24), *ρ*_2_ = 0.77 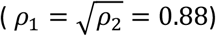 for short vs long term (*m* = 9), and *ρ*_1_ = 0.84 for medium vs long term (*m* = 34). This gives an overall weighted mean of *ρ*_1_ = 0.77.

Therefore, we chose to use *ρ*_1_ = 0.8 for both mean differences and change scores with one degree of separation. To cover the range of correlations observed, we chose to perform sensitivity analyses with *ρ*_1_ = 0.5 and *ρ*_1_ = 0.95.

### D Implementation of model in JAGS

We implemented our models in a Bayesian framework using JAGS [5]. We used four chains and assessed convergence by inspecting MCMC trace plots and using the Brooks-Gelman-Rubin 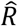 statistic [6, 7]. For all non SSVS models (see Section E) we assigned non-informative prior distributions to all parameters. For heterogeneity, we specified *τ*∼Unif(0,5). For the intercept and all regression coefficients we used *N*(0, 100^2^). An upper limit of 5 for heterogeneity and a standard deviation of 100 is large on the zBMI scale (which follows a standard normal in the general population). We used an adaptive phase of 10,000 iterations, a burn in of an additional 40,000, and a further 30,000 iterations from which we drew our posterior samples.

All code required to implement our analysis is provided in the GitHub repository here: https://github.com/AnnieDavies/Obesity_Synthesis

### E Selection of interactions via stepwise stochastic search variable selection (SSVS) models

#### E.1 SSVS model

To select interaction terms 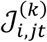, we use a Bayesian stochastic search variable selection (SSVS) model from Efthimiou et al [8]. While fitting the model, interactions are selected at each iteration of the MCMC using indicator variables *I*_*j*_ ∈ {0,1} in the prior distributions of the interaction coefficients *η*_*j*_. The priors are defined as the sum of two normal distributions. The value of the indicator variable at each iteration specifies which of these distributions to use. The first is very narrowly distributed around zero and is used when *I*_*j*_ = 0 (the *j*^*th*^ interaction term is not selected). The second is wider, allowing for non-zero values of the coefficient and is used when *I*_*j*_ = 1 (the *j*^*th*^ interaction is selected). By inspecting the posterior distribution of the indicator parameter for each interaction term *j* we can assess the proportion of times the interaction was selected. The more important an interaction, the more often it will be selected.

The SSVS version of our model is defined in the same way as in Section B but with the following prior distributions on the interaction coefficients,

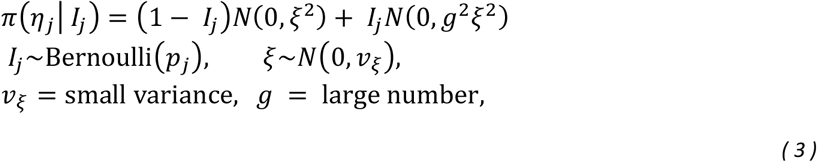

where *j* = 1, … , *l* and *l* is the number of interactions from which to select. For all other model parameters we use the non-informative prior distributions described in Section D. The variance *v*_*ξ*_, which specifies the width of the narrow distribution, should be such that it only allows for effects that are `practically zero’ relative to the scale of the data. The parameter *g*, which specifies the width of the wider distribution, should be large enough to allow for non-zero (but sensible) values of the parameter. Informed by [8], the scale of zBMI, and preliminary tests on our data, we chose *v*_ξ_ = 10^−3^ and *g*^2^ = 100.

In Equation ( 3 ), the indicator variables are assigned Bernoulli prior distributions with probabilities *p*_*j*_. These probabilities can be used to define informative priors based on our beliefs about the importance of interaction terms. Otherwise, setting *p*_*j*_ = 0.5 ∀*j* defines non-informative (equi-probable) priors which choose the interactions based on information from the data alone. In our step-wise procedure described in Section E.2 we use the latter.

#### E.2 Step-wise SSVS procedure

As described in the main paper, we selected interaction terms for our model by applying the SSVS method in a step-wise process. Each step involved a different set of interaction terms from which to select. In the following we list the model specified at each step and give the implementation details used to fit it in JAGS [5].

i. Step 1: Model with no interactions
  - We used an adaptive phase of *n*_adapt_ = 10,000 iterations, an additional burn in of *n*_burn_ = 20,000 , followed by a further *n*_samp_ = 30,000 iterations from which we drew our posterior samples.
ii. Step 2: Model with interactions between age and all other indicator variables
  - Implementation details: *n*_adapt_ = 10,000, *n*_burn_ = 20,000, *n*_samp_ = 30,000.
iii. Step 3: Model with interactions between ‘change of behaviour targeted’ (diet and/or physical activity) and all other indicators
  - As described in Section 3.1 of the main paper, behaviour targeted was coded as two binary variables indicating whether the intervention targeted physical activity alone or a combination of diet and physical activity. An intervention targeting diet alone is then indicated by setting both these variables to zero.
  - This model included interactions between each of these variables (physical activity alone and diet & physical activity) and every other indicator.
  - Implementation details: *n*_adap*t*_ = 10,000, *n*_burn_ = 40,000, *n*_samp_ = 30,000.

To define our final model, we chose interaction terms that were selected more than 50% of the time in steps 2 and 3. We performed steps 1-3 separately for the random effects and fixed effect models.

#### E.3 Deviations from our analysis plan (indicator selection)

In our analysis plan we set out to use the SSVS method to select intervention indicators as well as interactions. However, the first two steps of our selection process (assessing collinearity and the proportion of identical responses) led to a manageable number of indicators (see Table 1 in the main paper) all of which were deemed important by our stakeholders. Therefore, we did not require the SSVS method to reduce the number of indicators further.

Our analysis plan also states that we would to assign informative probabilities to the SSVS prior distributions, describing the interactions that our stakeholders considered most and least probable. However, beyond expressing interest in interactions between age and behaviour targeted, our stakeholders encountered challenges in pinpointing particular interactions of interest. Additionally, preliminary analyses revealed that the use of informative priors led to the dominance of predetermined interactions, undermining our objective to select interactions based on the data.

### F Adjusting the intercept

In Section 3.2.3 of the main manuscript we evaluated our primary model for every possible combination of indicator variables. Each combination involved inserting values of 0 or 1 for each indicator. However, the model intercept was calculated using indicators centred at their mean value. Therefore, to interpret the outcome of the model as a mean difference in change from baseline in zBMI we need to convert the intercept from the centred model to the intercept for the non-centred model.

Using the notation from Section B, the fixed effects of the non-centred (*nc*) model (for control comparison studies) is

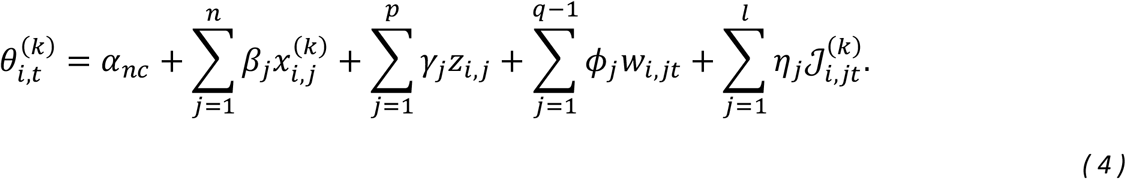

The equivalent model with centred (*c*) indicators is

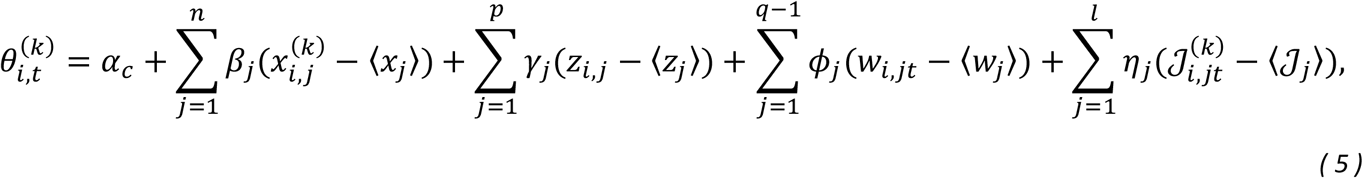

where the angular brackets ⟨ . ⟩ denote the mean value of that indicator (or interaction) over all observations in the data set. The intercept reported in the main paper is an estimate of the centred intercept, α_*c*_. To convert this to the non-centred intercep*t α*_*nc*_ we equate right hand side of Equations ( 4 ) and ( 5 ), which leads to

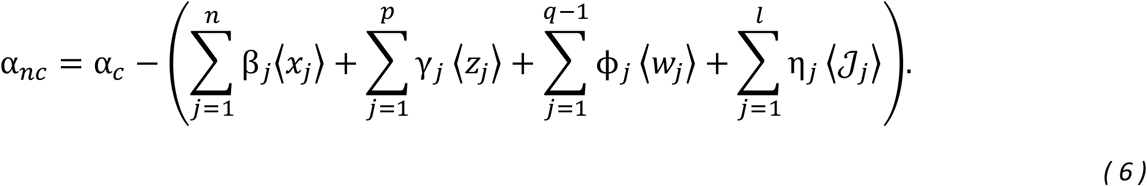

To obtain the predicted mean differences in Table 2 in the main paper, we calculated the intercept from Equation ( 6 ) using the mean values of each indicator variable used to centre the data, and the estimated intercept (α_*c*_) and regression coefficients from our primary analysis. This gave *α*_*nc*_ = −0.0003, which represents the effect of an intervention whose indicators are all equal to zero.

### G Study characteristics

**Table S1.**
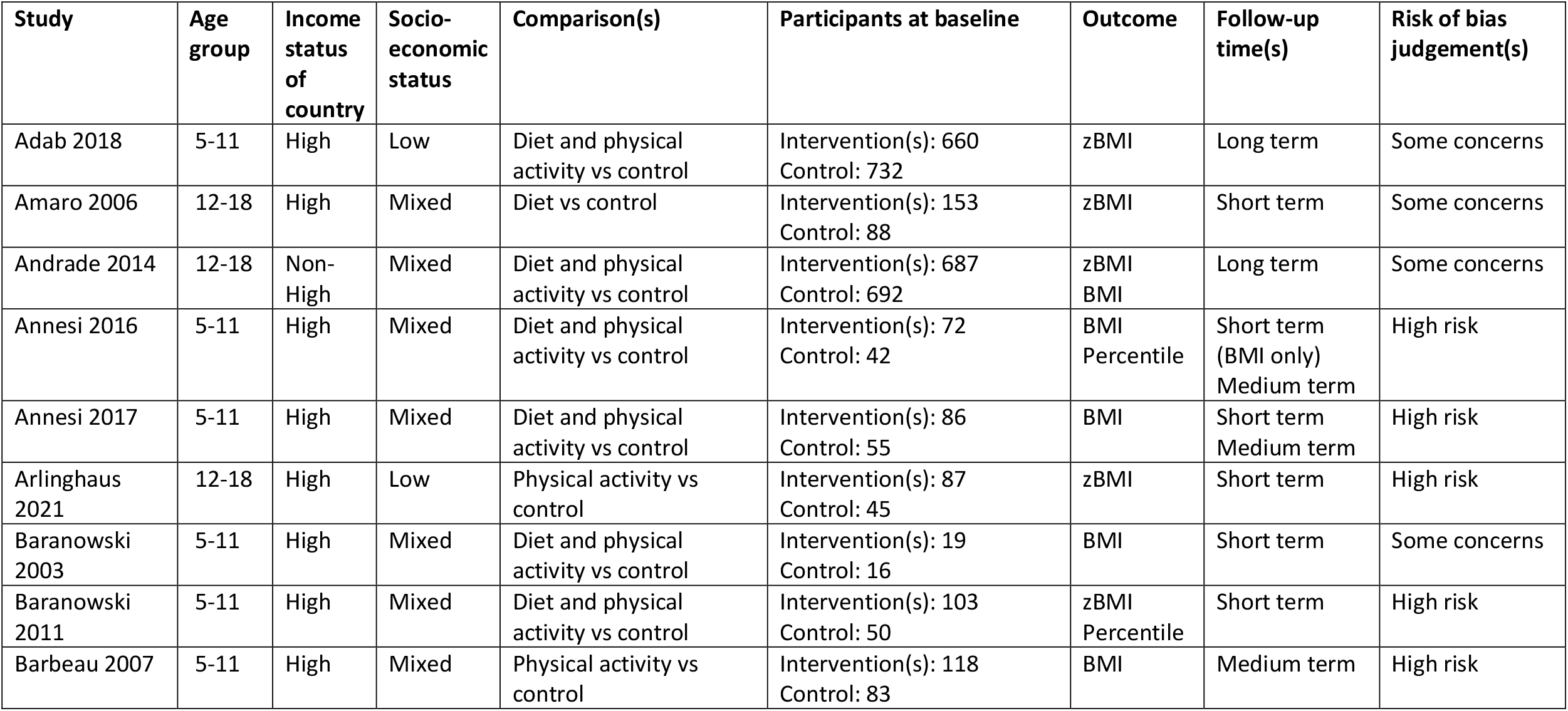
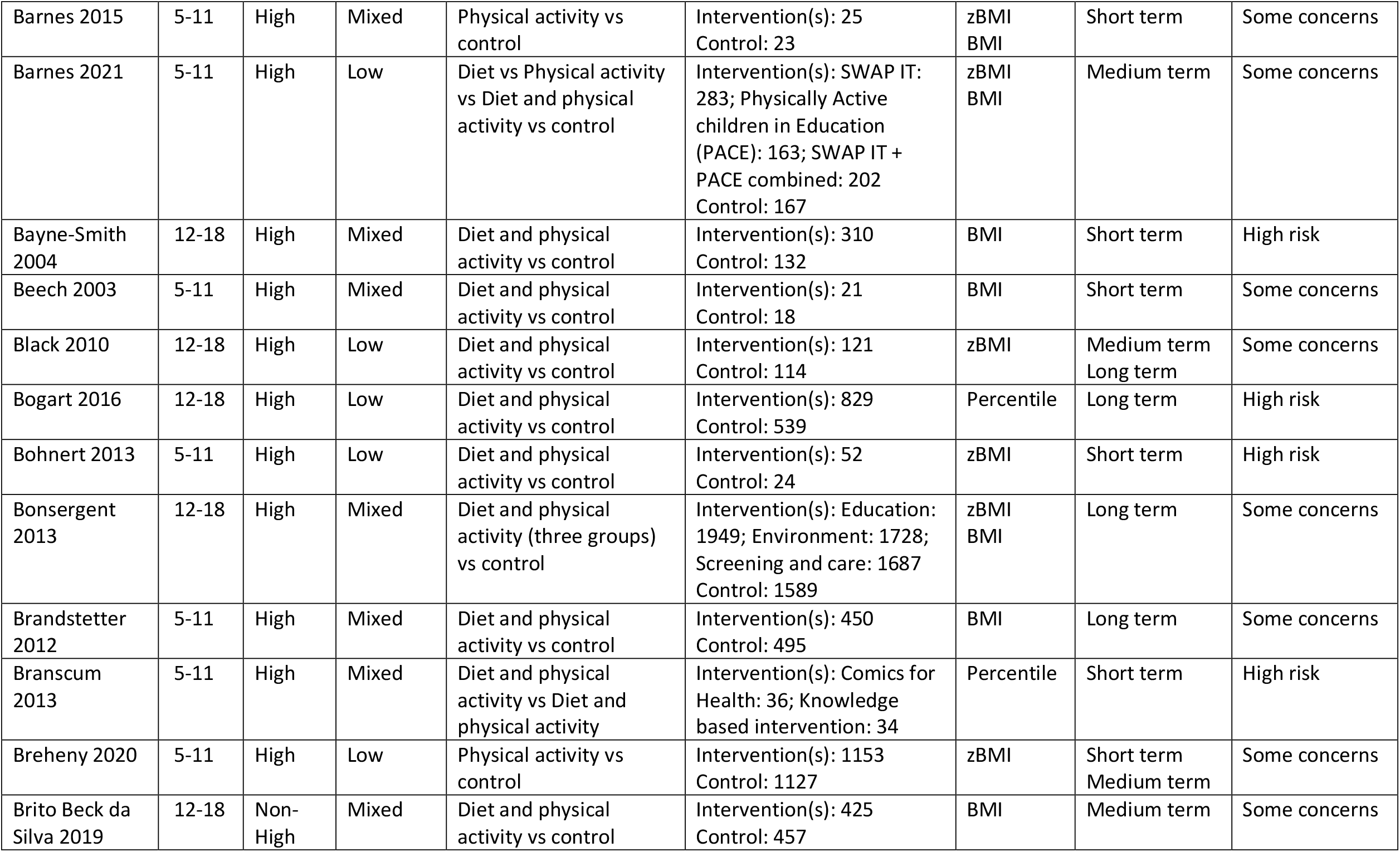
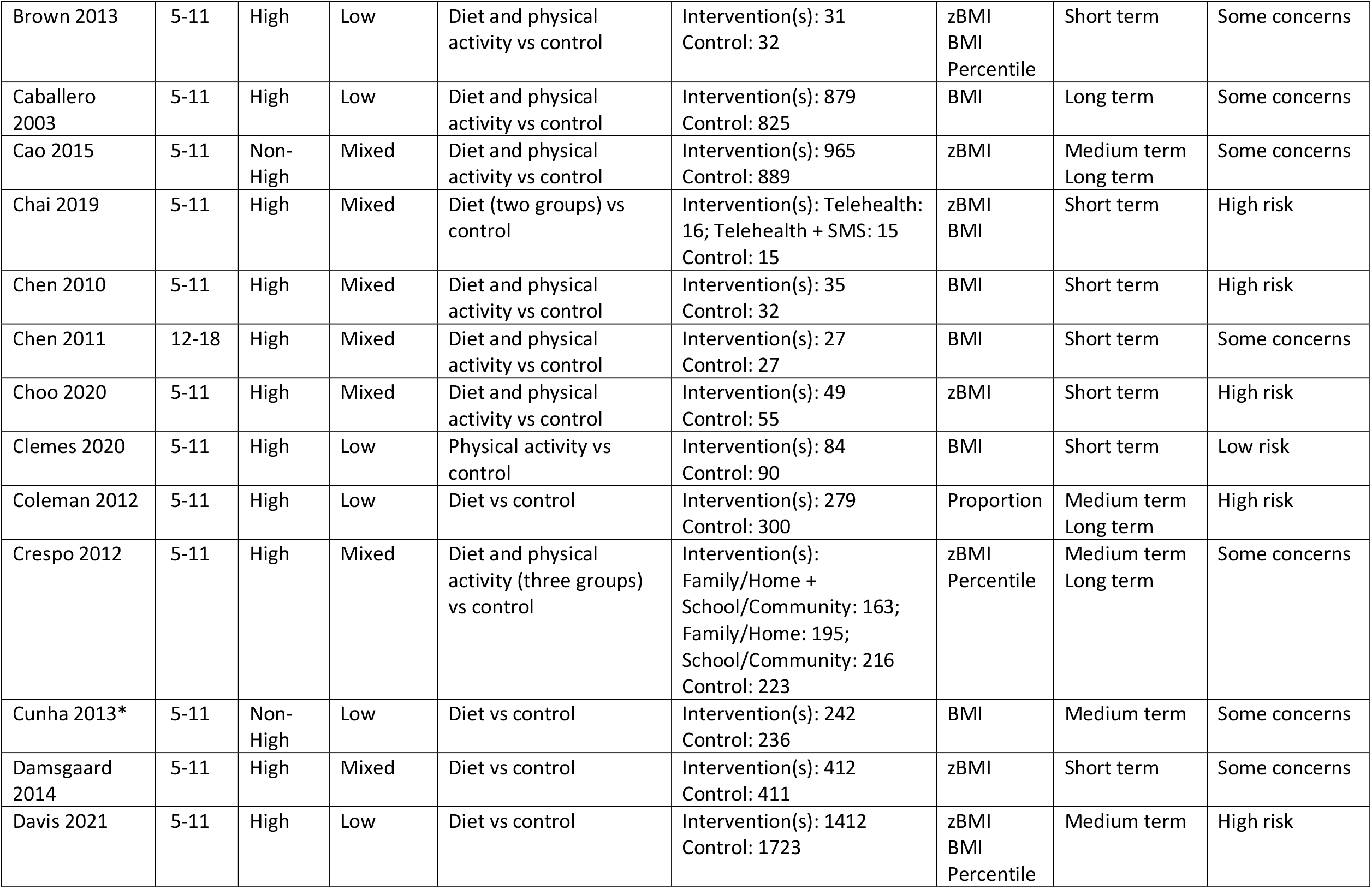
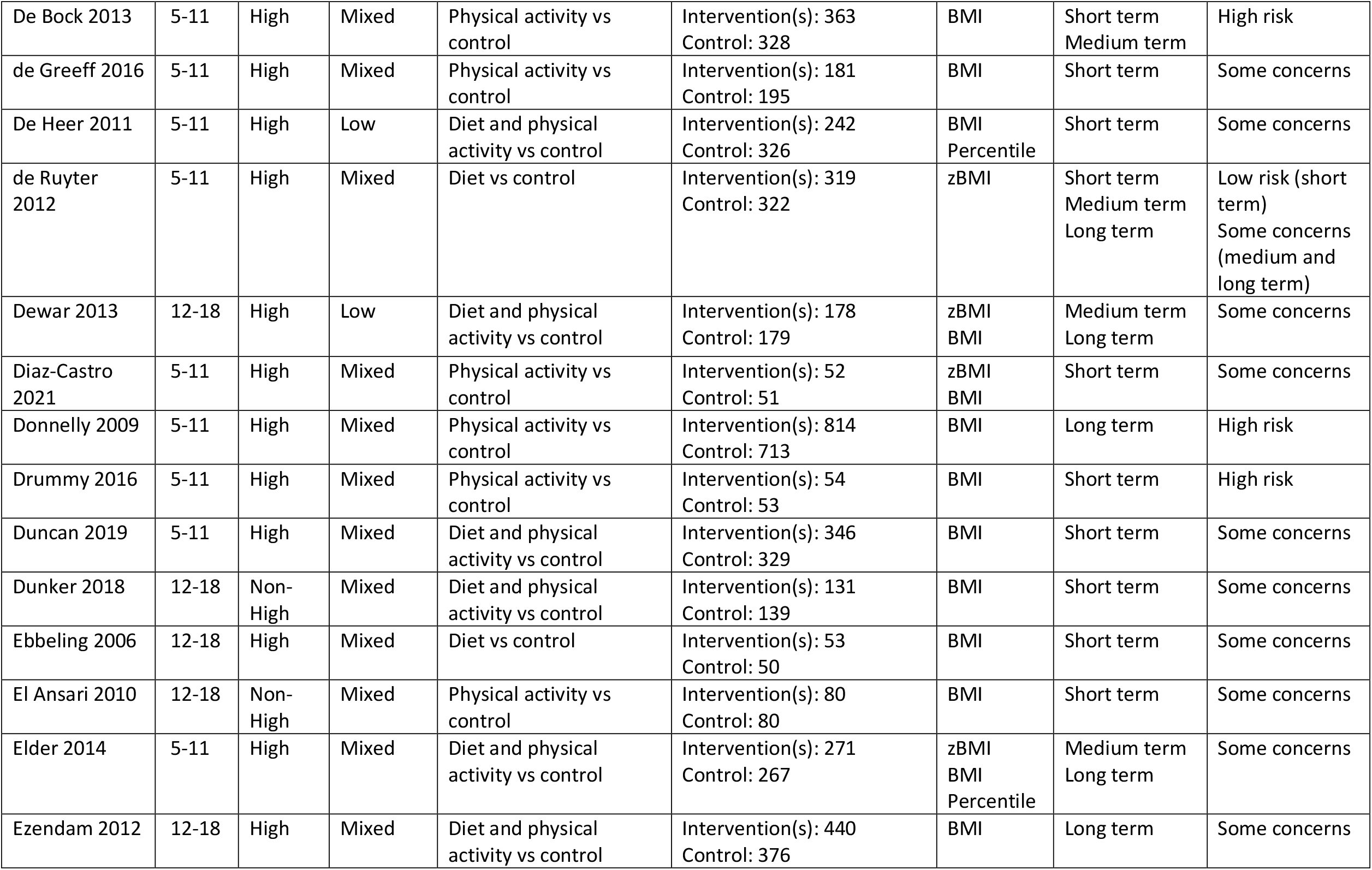
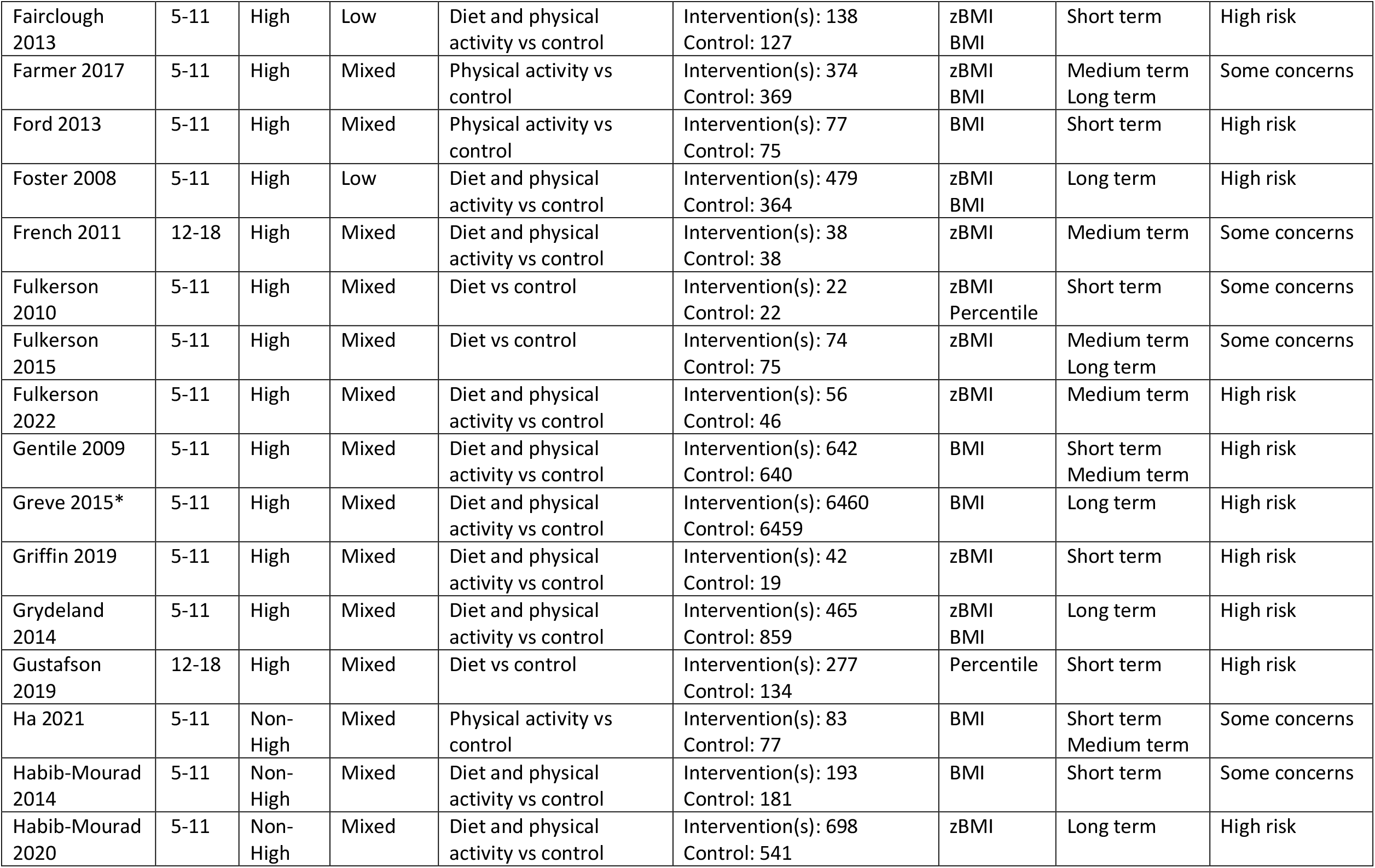
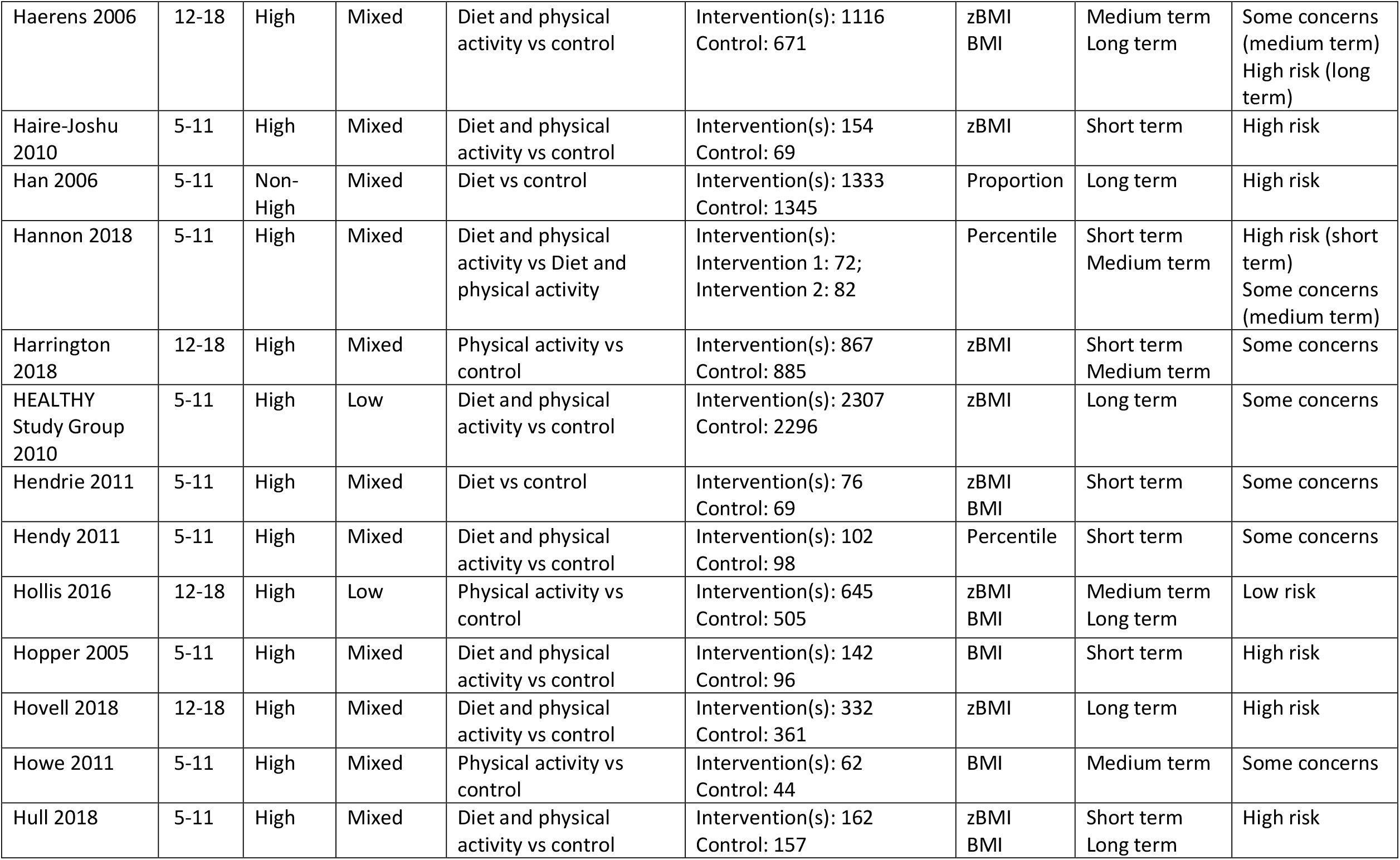
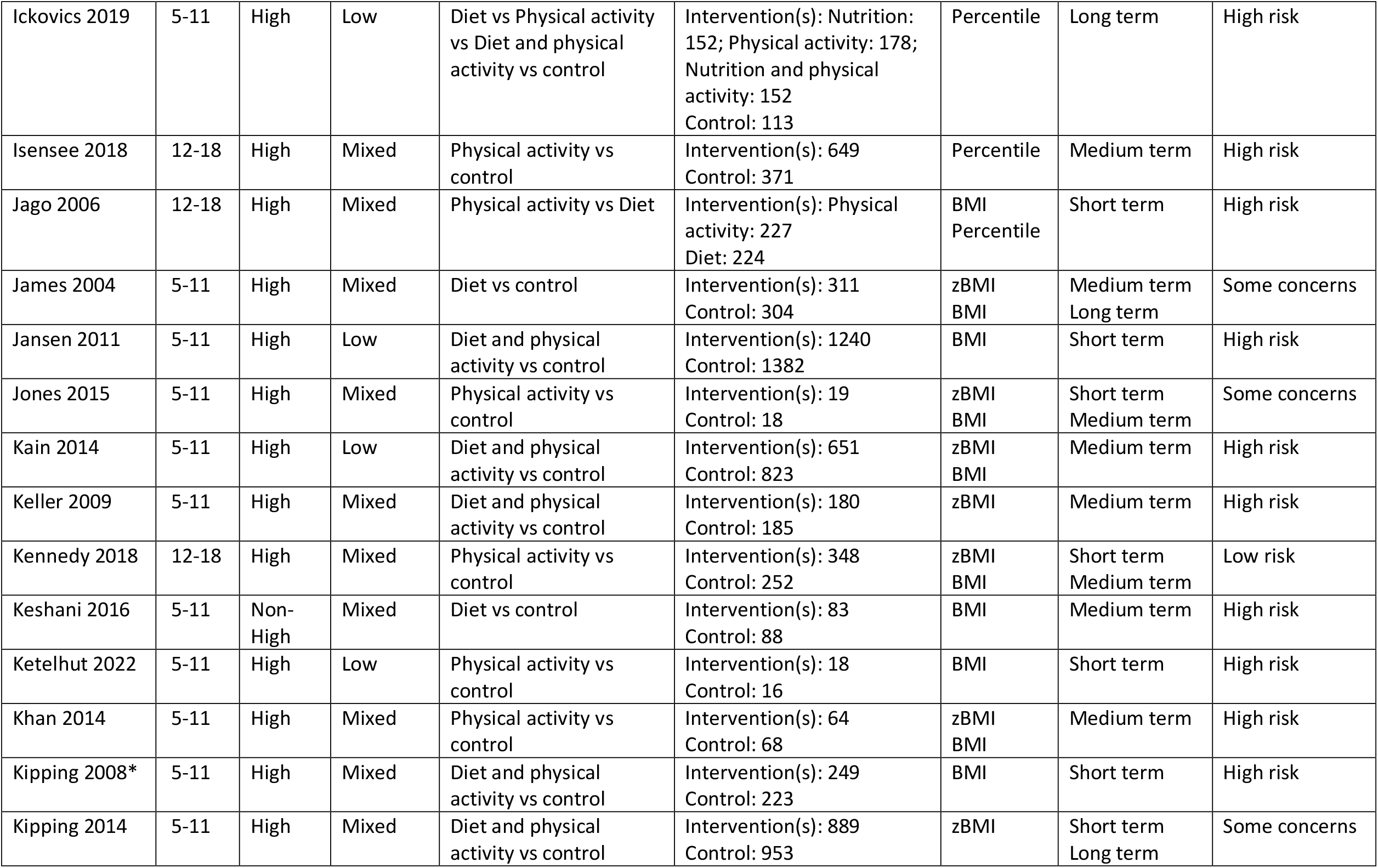
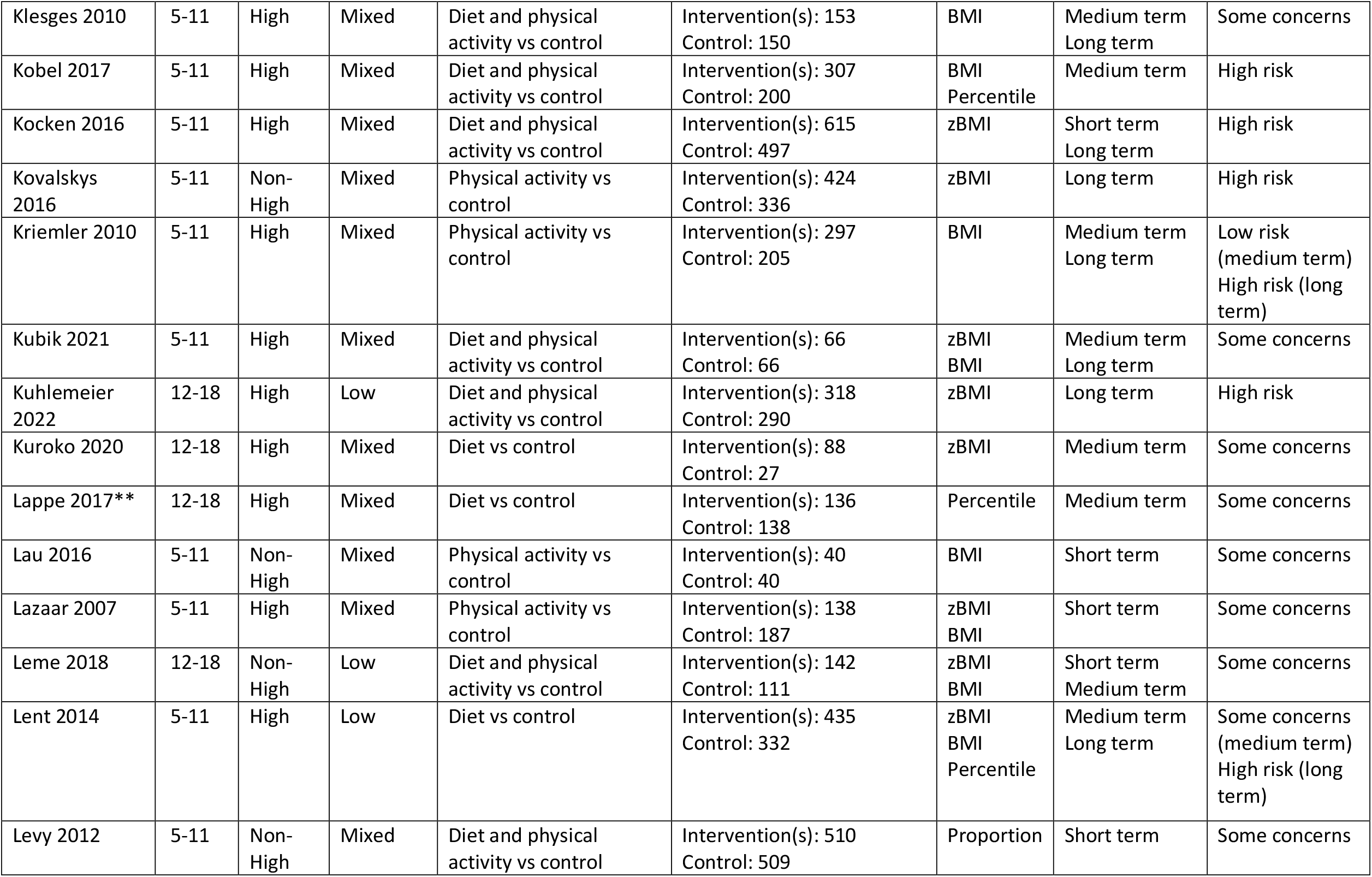
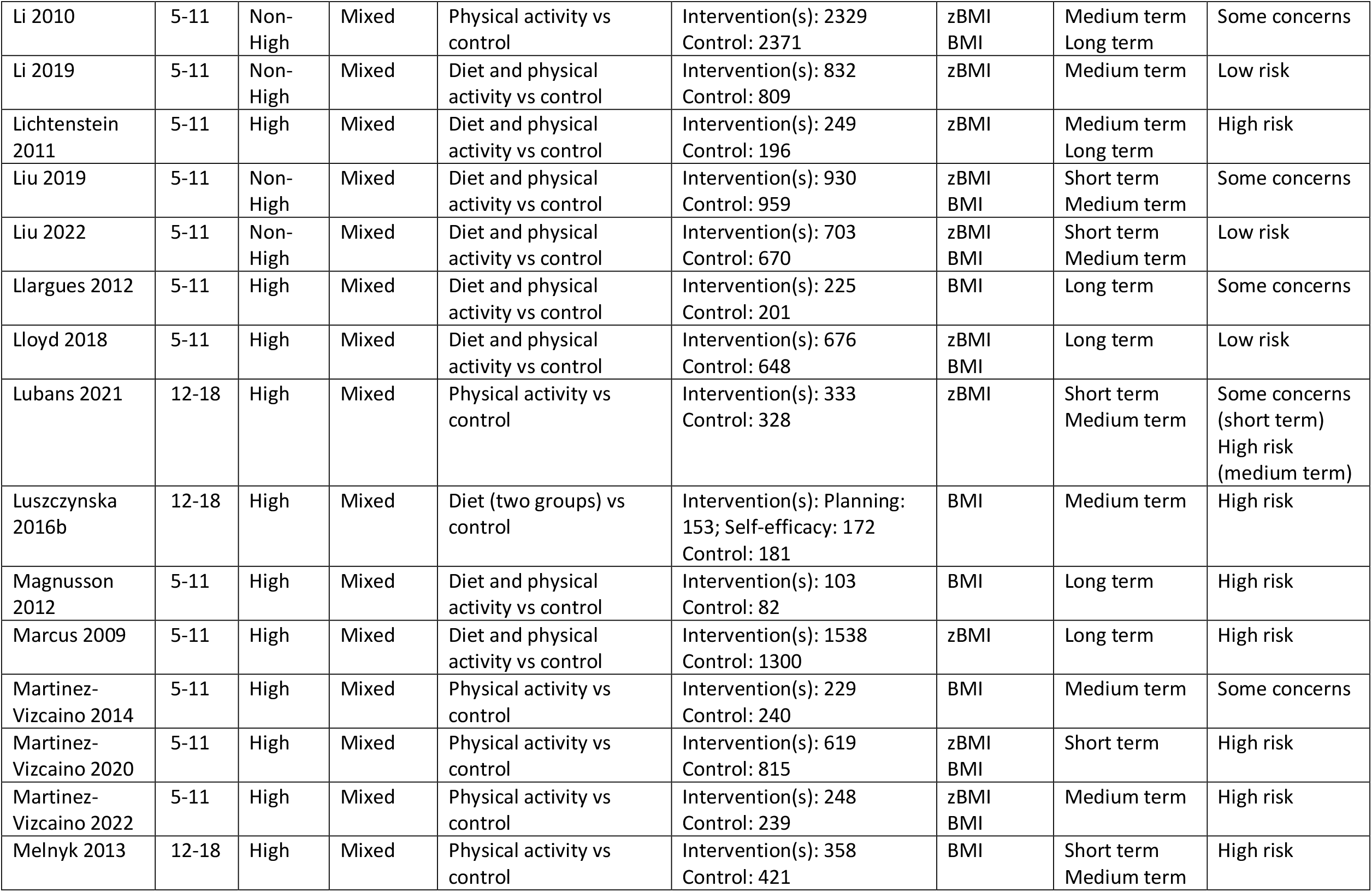
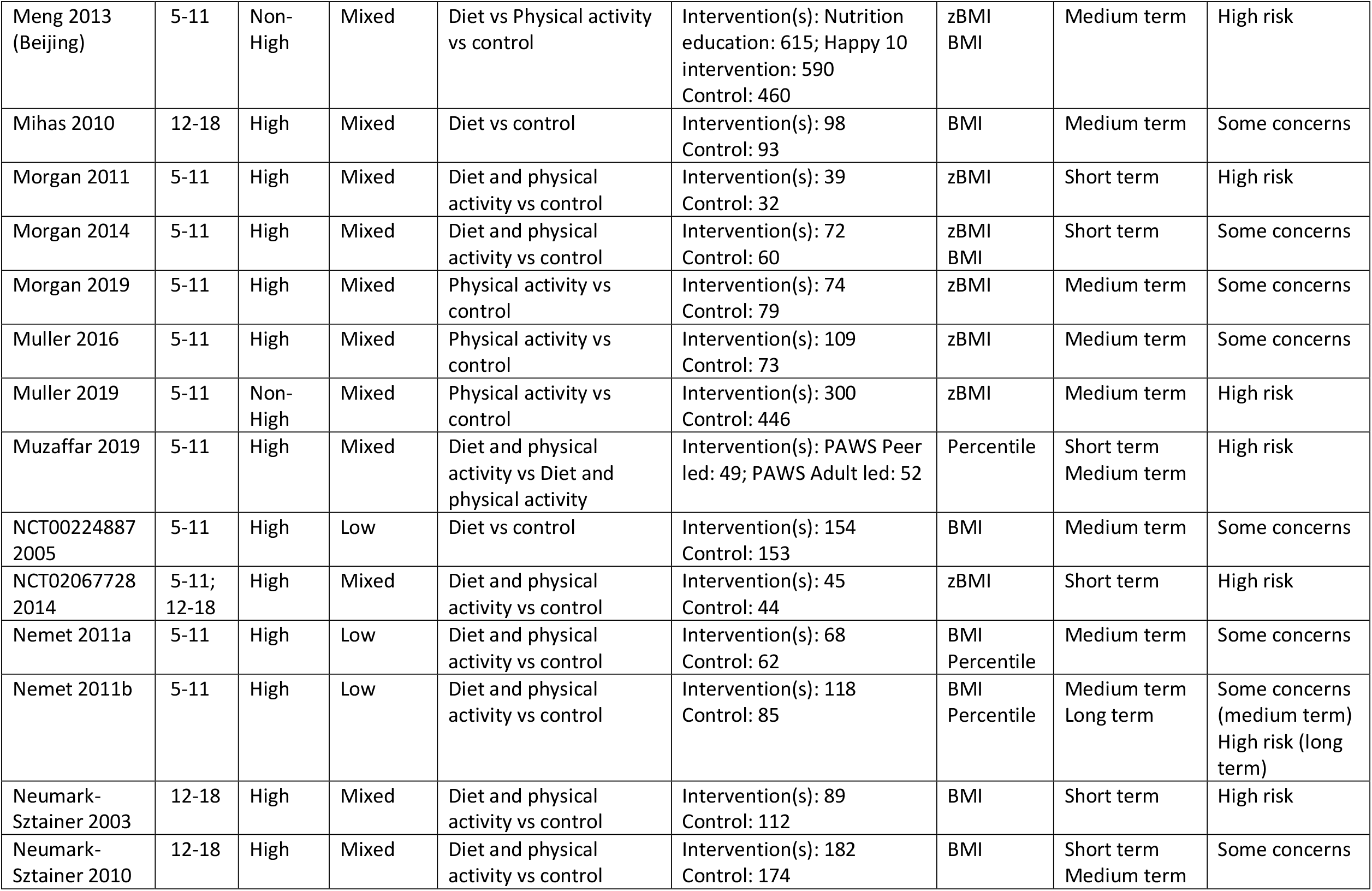
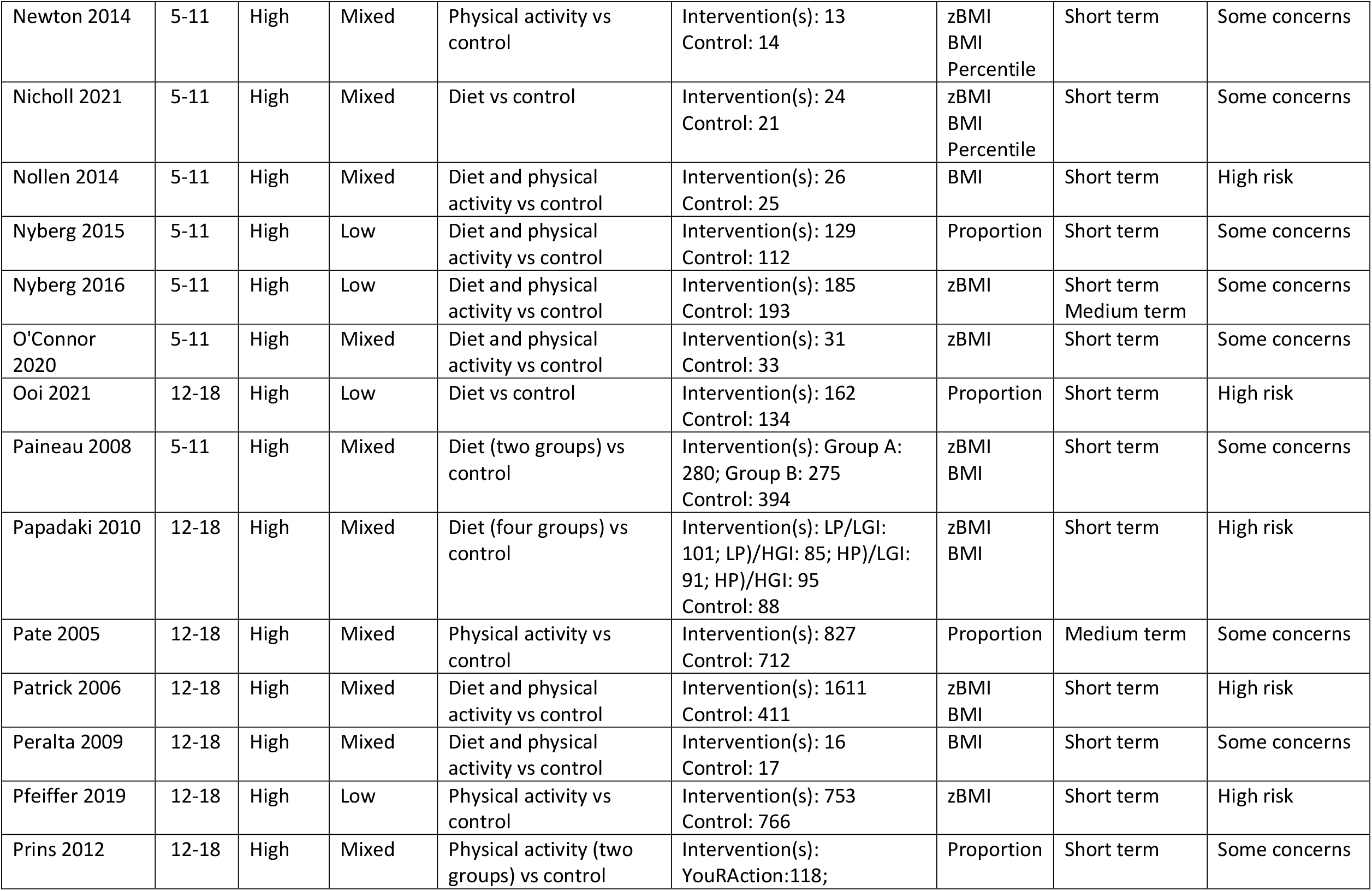
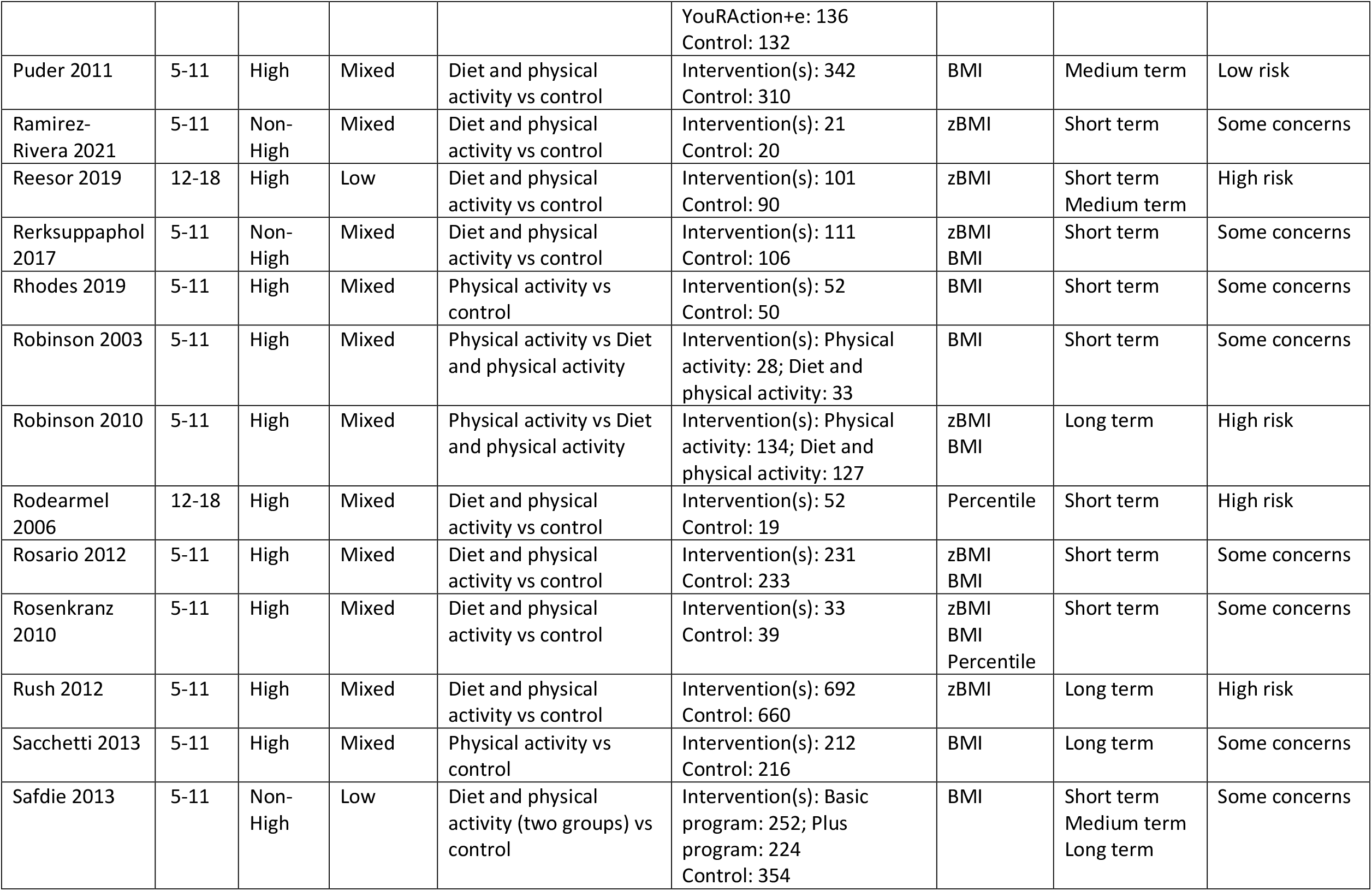
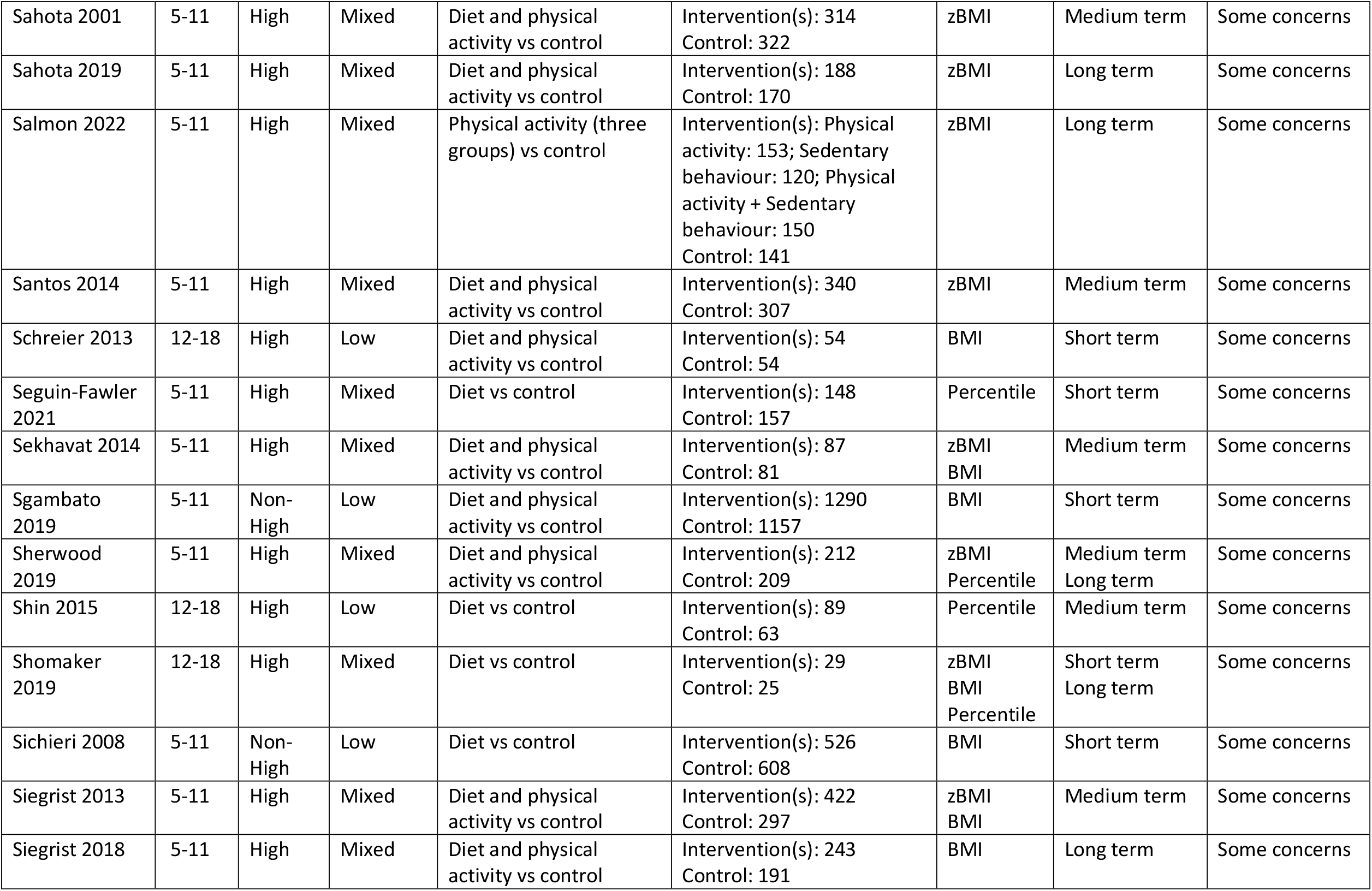
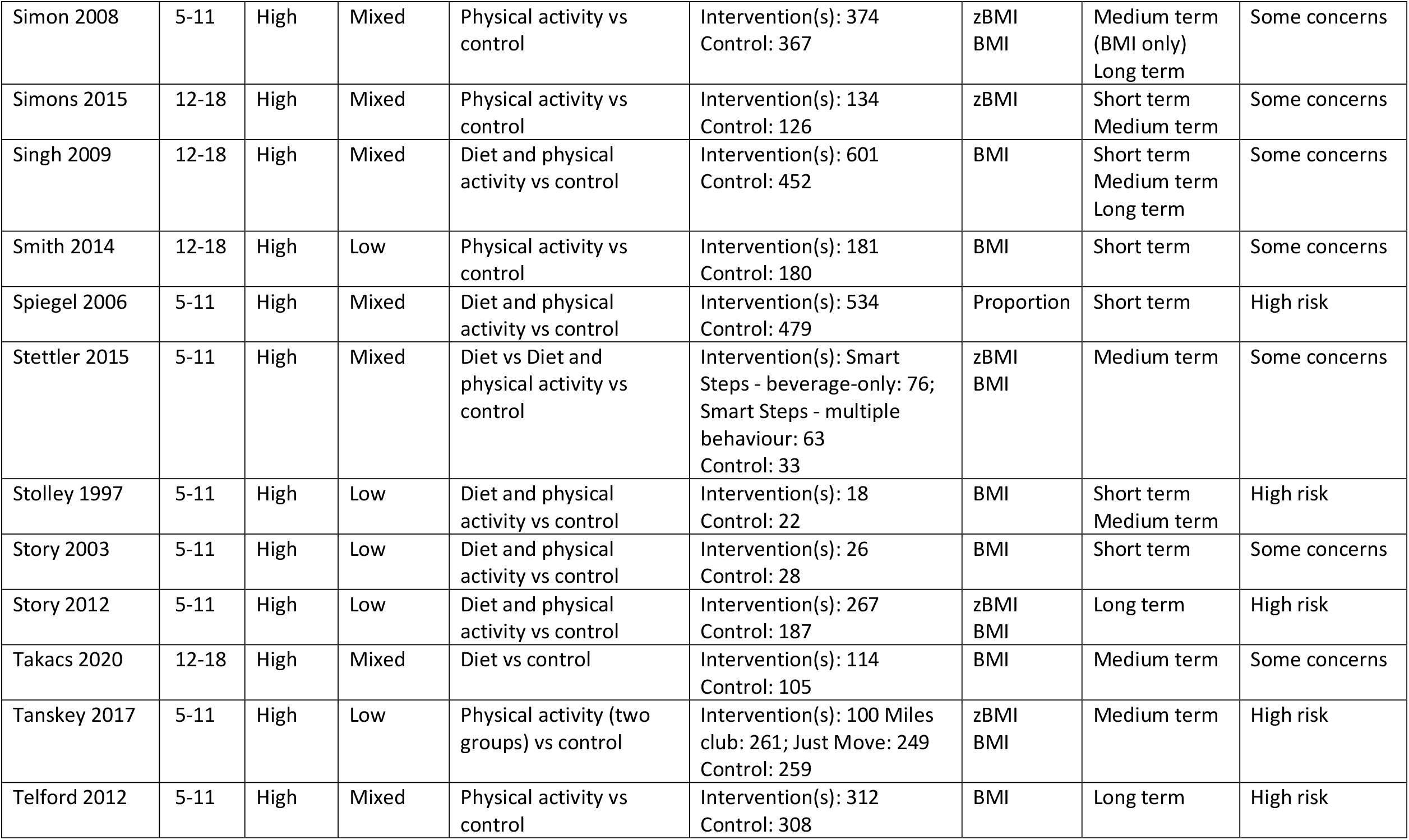
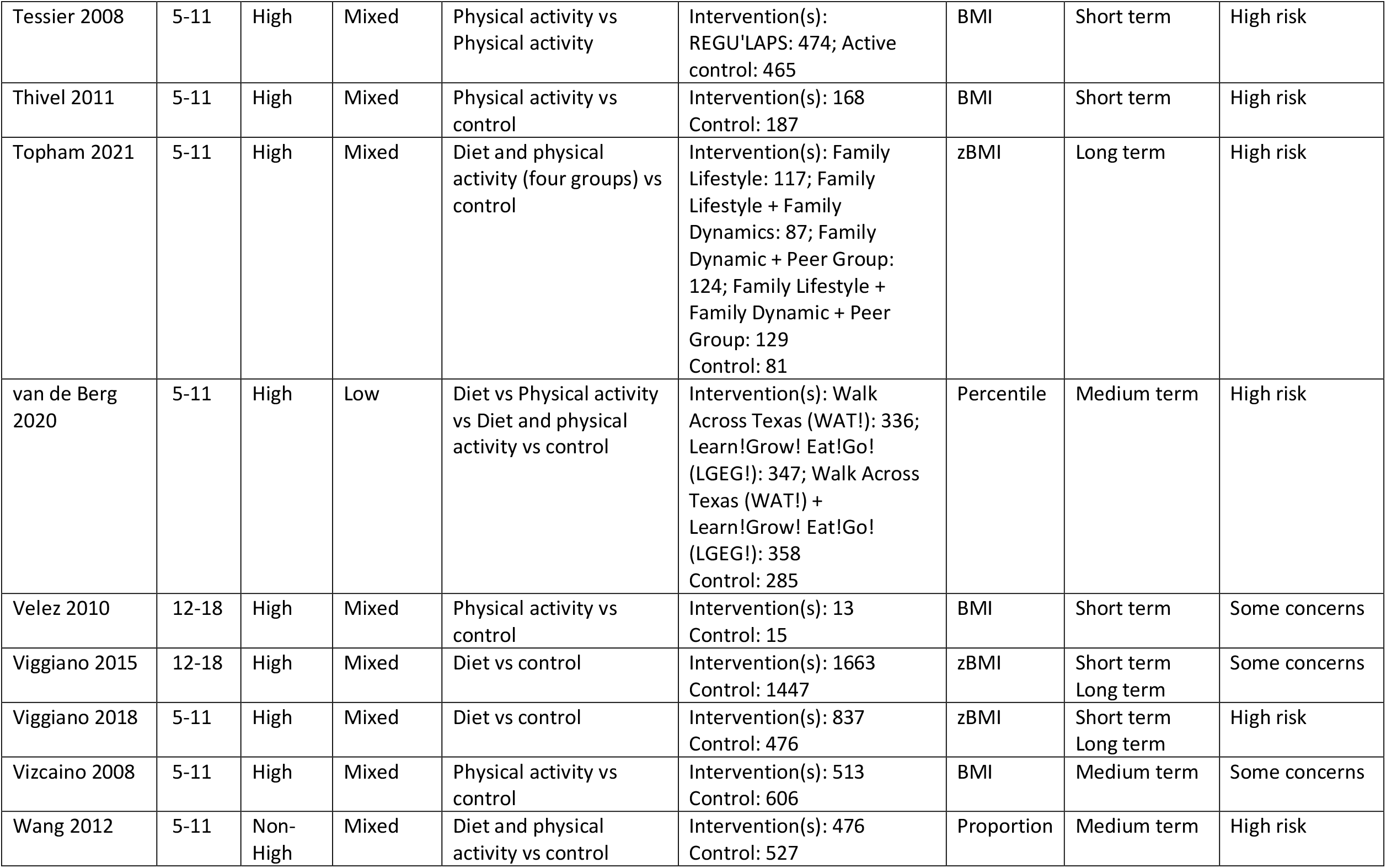
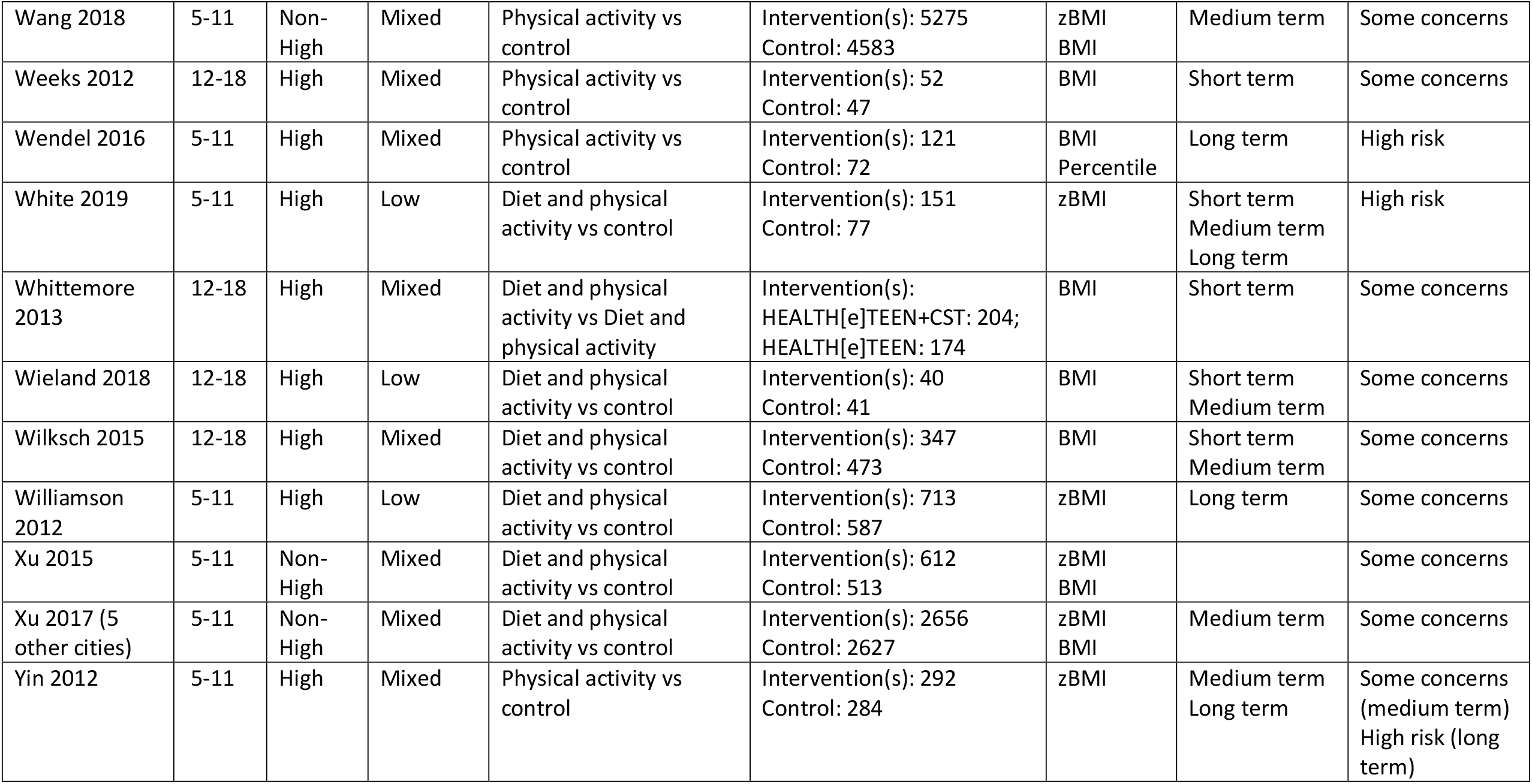
Summary of the characteristics of the trials included in our analyses. For studies that provide data on multiple outcomes, we selected data in the following order of preference: (i) zBMI, (ii) percentile (mapped), (iii) BMI (mapped). The BMI-only sensitivity analysis included all studies that provided data on BMI. *\*Studies that only provide contrast-level data on BMI and could not be mapped to zBMI. These studies were excluded from the main analysis but were included in the BMI-only sensitivity analysis*. *\*\*Study that only provides contrast-level data on percentile and could not be mapped to zBMI. This study was excluded from all analyses*.

### H Selecting indicator variables

The correlation matrix between the intervention indicators is shown in **Figure S*1***. We observed two absolute correlations ≥ 0.5 between (i) the total duration of the intervention and the duration of the peak engagement period (correlation of 0.66), and (ii) school setting and community setting (correlation of -0.72). Based on (i) we chose to remove peak duration from our set of indicators. Correlation (ii) reflects the fact that interventions tended to be based in either a school or a community environment, while home elements were often in supplement to one of these settings. Since the effect of all settings were deemed of interest by our stakeholders, we did not remove either of these variables.

**Figure S*2*** shows the proportion of interventions that were coded as 1 (rather than 0) for each intervention indicator variable. We observed six indicators that yielded identical responses more than 80% of the time: delivered in the home (81.6% No), targets activity (81.2% Yes), applied continuously (96.5% Yes), delivered to the child as part of a group (82.7% Yes), involved a single phase (81.2% Yes), commercial interests involved (89.4% No). Based on these results, we chose to remove the continuous, single phase and commercial interests indicators due to a lack of information. Next, we re-defined the home indicator to also include `home activity’. This meant that home was coded as 1 if the intervention was delivered in the home OR included some home based activity for the child. We also re-coded the group and individual indicators as one variable coded as 1 if the intervention was delivered to the child individually (either exclusively or in combination with group delivery) and 0 if there was no individual element. Finally, we chose to combine the flexibility and choice indicators which, while not quite reaching our 80% threshold, were both mostly coded as 0 (No) and represent a similar concept (i.e. the adaptability of the intervention to the preferences of the recipient or deliverer of the intervention). Since evaluating the effect of activity based interventions is of particular interest, we did not remove this indicator.

**Figure S1:**
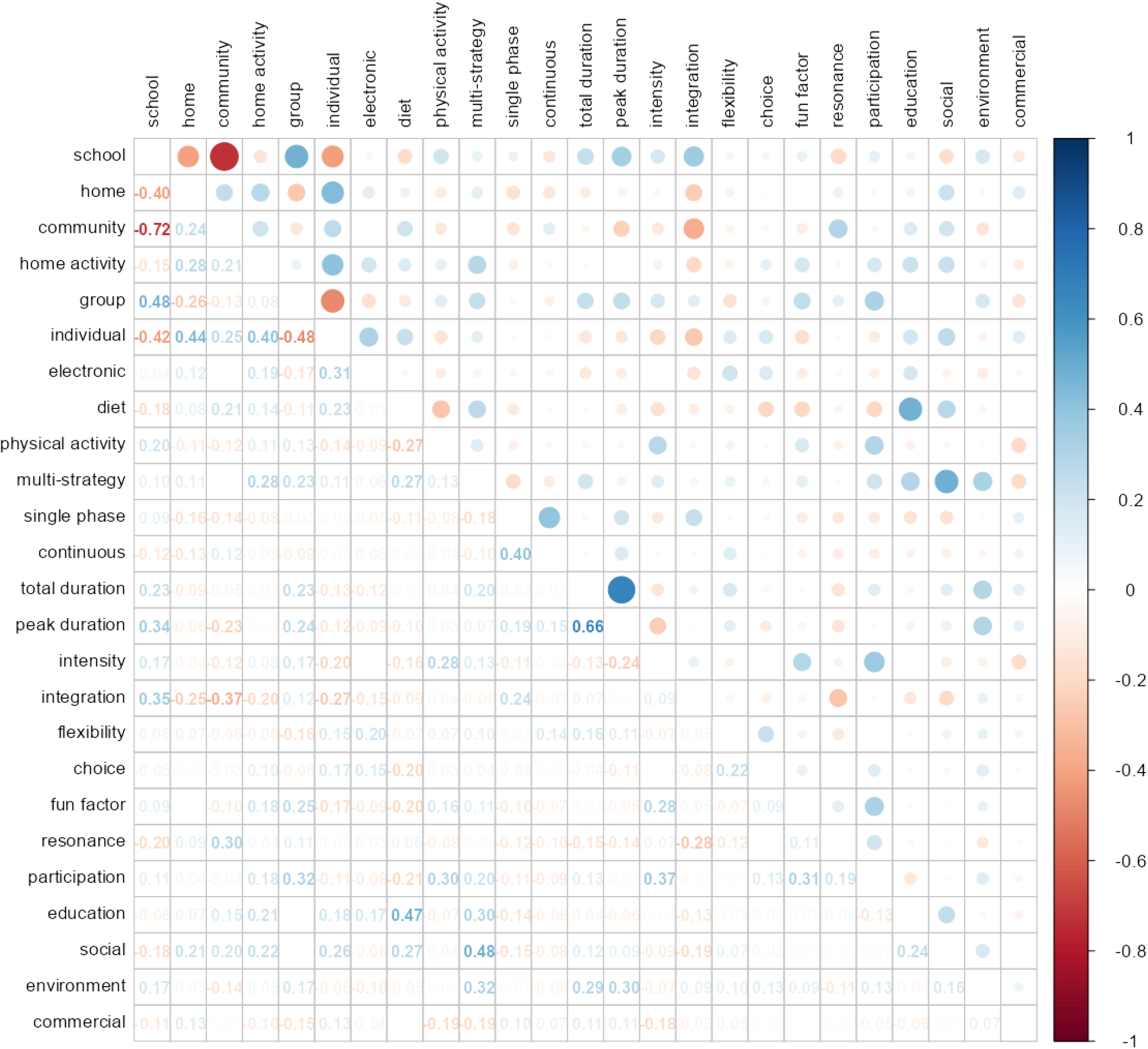
Correlation matrix describing the collinearity between pairs of intervention indicators. The colour and size of circles in the upper half of the matrix indicates the correlation between those variables. Negative correlations are indicated in red and positive correlations in blue (as shown in the key on the right hand side). The value of the correlation is shown in the equivalent box in the lower half of the matrix. The lowest correlations are very faint and sometimes cannot be seen.

**Figure S2:**
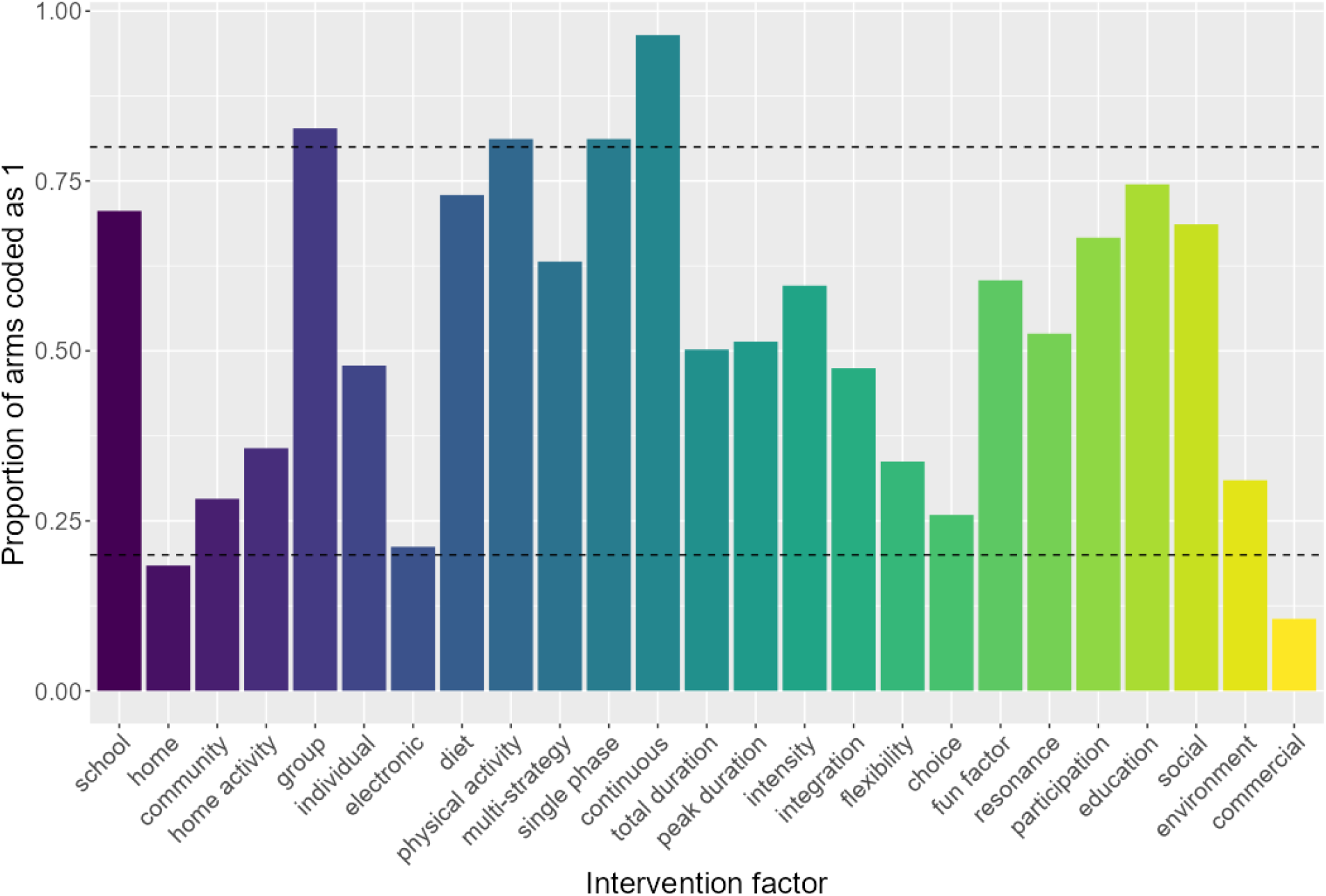
Bar chart showing the proportion of identical responses for each intervention indicator. Each bar represents the proportion of intervention arms coded as 1 in the data set (see Section A for a description of each indicator and its coding). The horizontal dotted lines are at 0.2 and 0.8 representing our preliminary cut off of 80% identical responses (for 0 and 1 respectively).

### I Selecting interactions via SSVS

The results of the SSVS models at each step are shown in Sections I.1 and I.2 for the primary (random effects) and secondary (fixed effects) analyses respectively. **Table S*2*** compares the fit of the different models using the deviance information criterion (DIC) [9]. For both the RE and FE procedures, the final model with selected interactions has the lowest DIC indicating the best fit. The RE models have lower DIC (better fit) than the FE models.

**Table S2.**
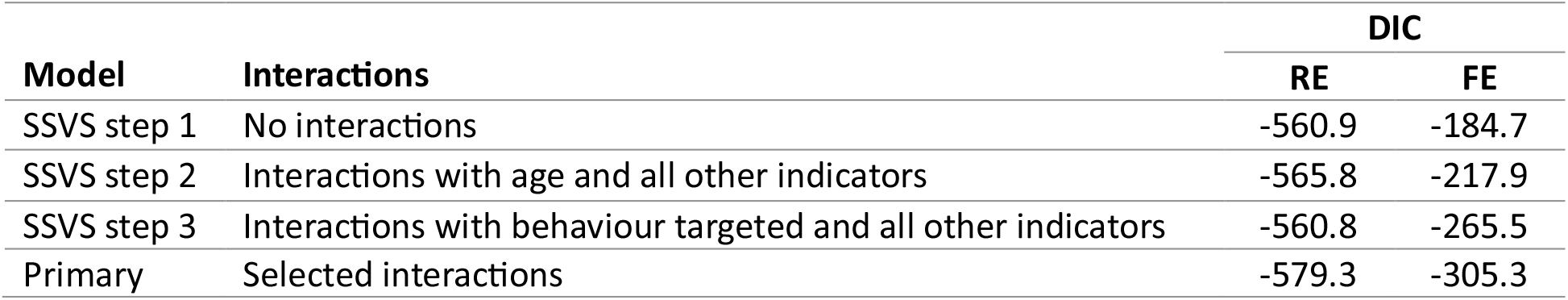
Deviance information criterion (DIC) for the models fitted at each step of the SSVS procedure (for both random effects and fixed effects).

#### I.1 Primary analysis (random effects)

##### I.1.1 Step 1: No interactions

**Figure S3:**
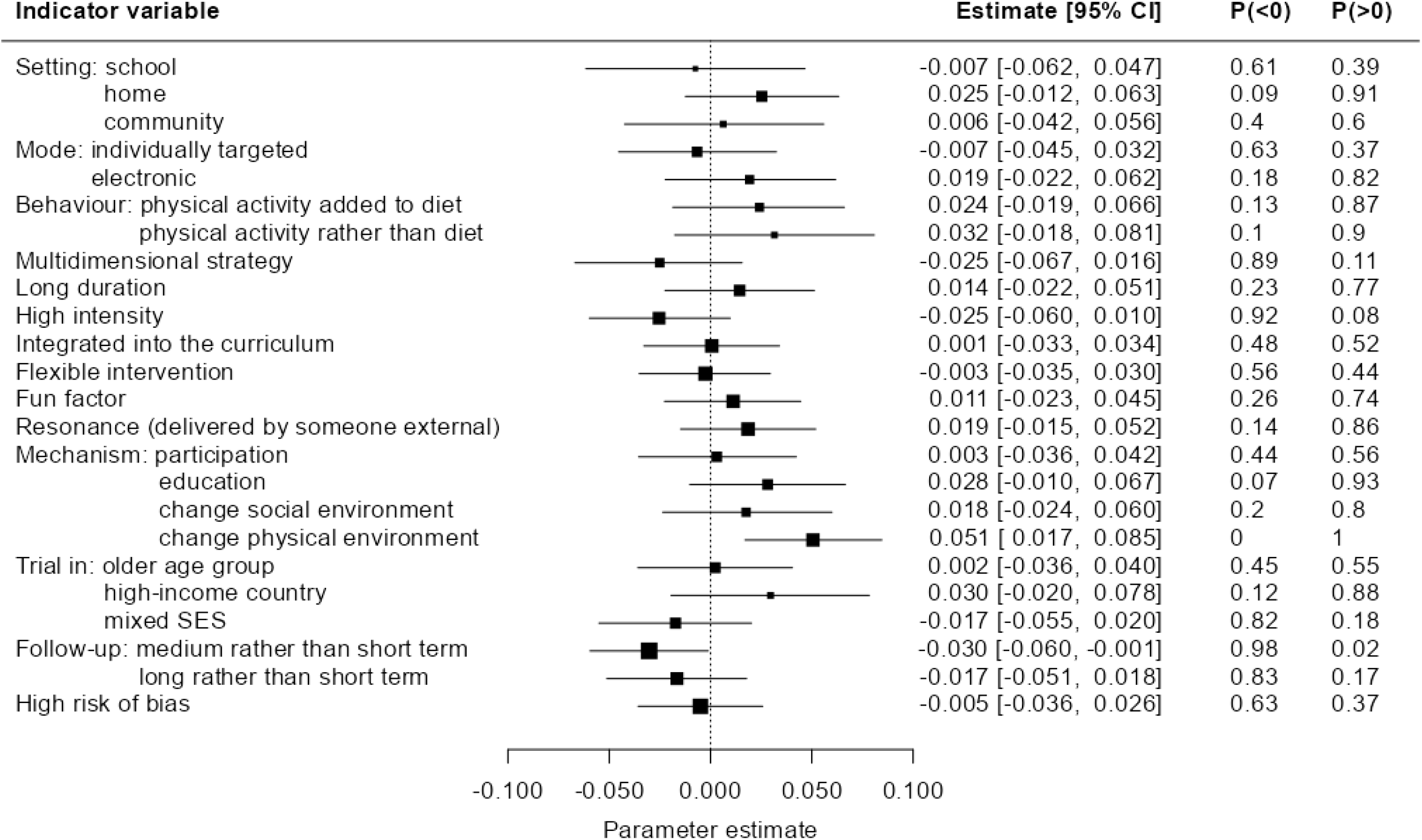
Parameter estimates from step 1 of the SSVS procedure (model with no interactions) for the primary (random effects) model. We list the probability that each coefficient is less than or greater than zero, *P*(< 0) and *P*(>0). The estimates of the intercept and heterogeneity parameter are *α* [95% *CI*] = −0.039[−0.054, −0.023] and *τ* [95% *CI*] = 0.085 [0.072, 0.099] respectively.

##### I.1.2 Step 2: Interactions with age

**Figure S4:**
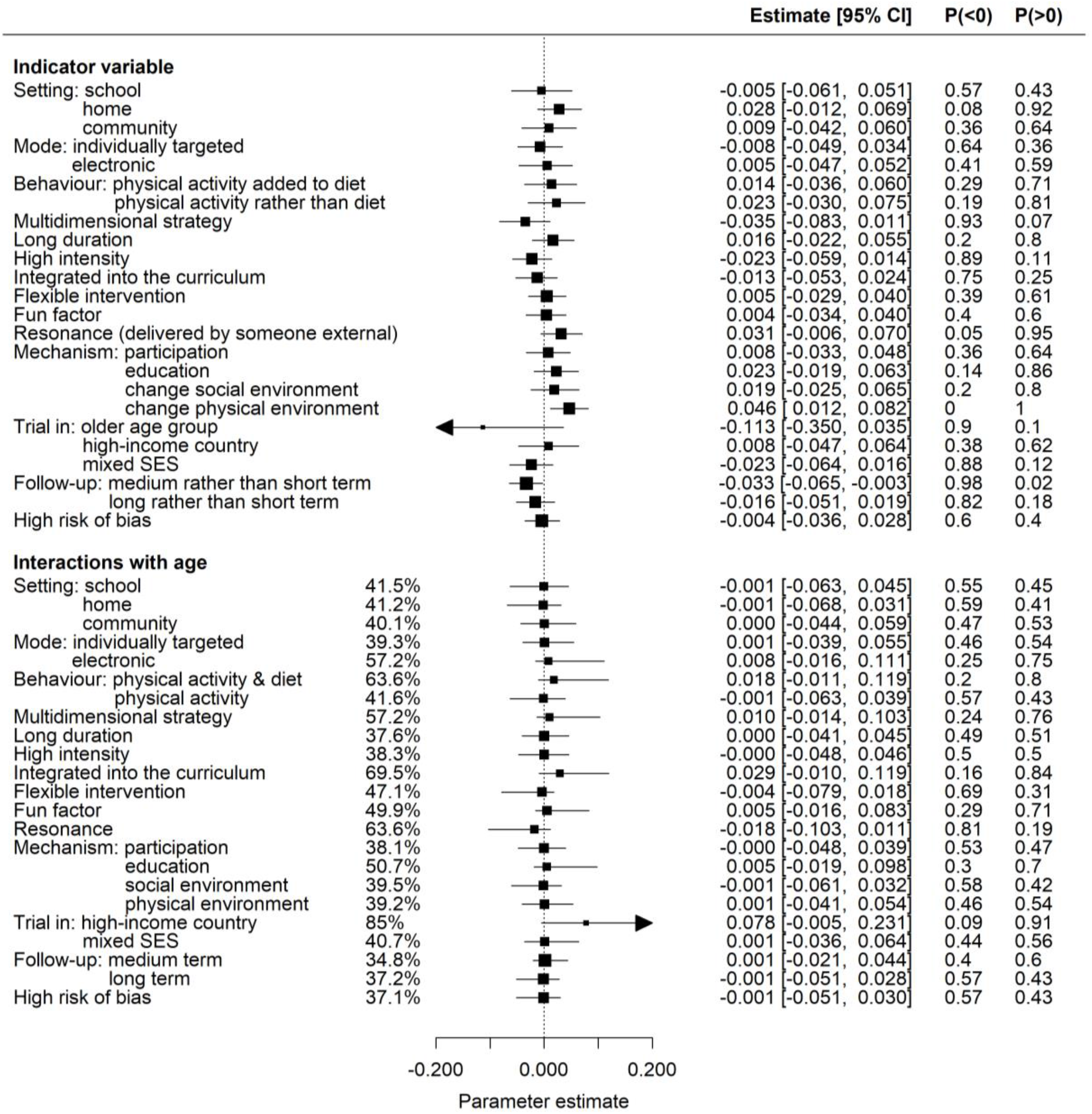
Parameter estimates from step 2 of the SSVS procedure (interactions between age and all other indicators) for the primary (random effects) model. We list the probability that each coefficient is less than or greater than zero, *P*(< 0) and *P*(> 0). For each interaction term we list the percentage of times it was selected by the SSVS model. The estimates of the intercept and heterogeneity parameter are *α* [95% *CI*] = −0.038 [−0.053, −0.023] and *τ* [95% *CI*] = 0.082 [0.070, 0.097] respectively.

##### I.1.3 Step 3: Interactions with change in behaviour targeted

**Figure S5:**
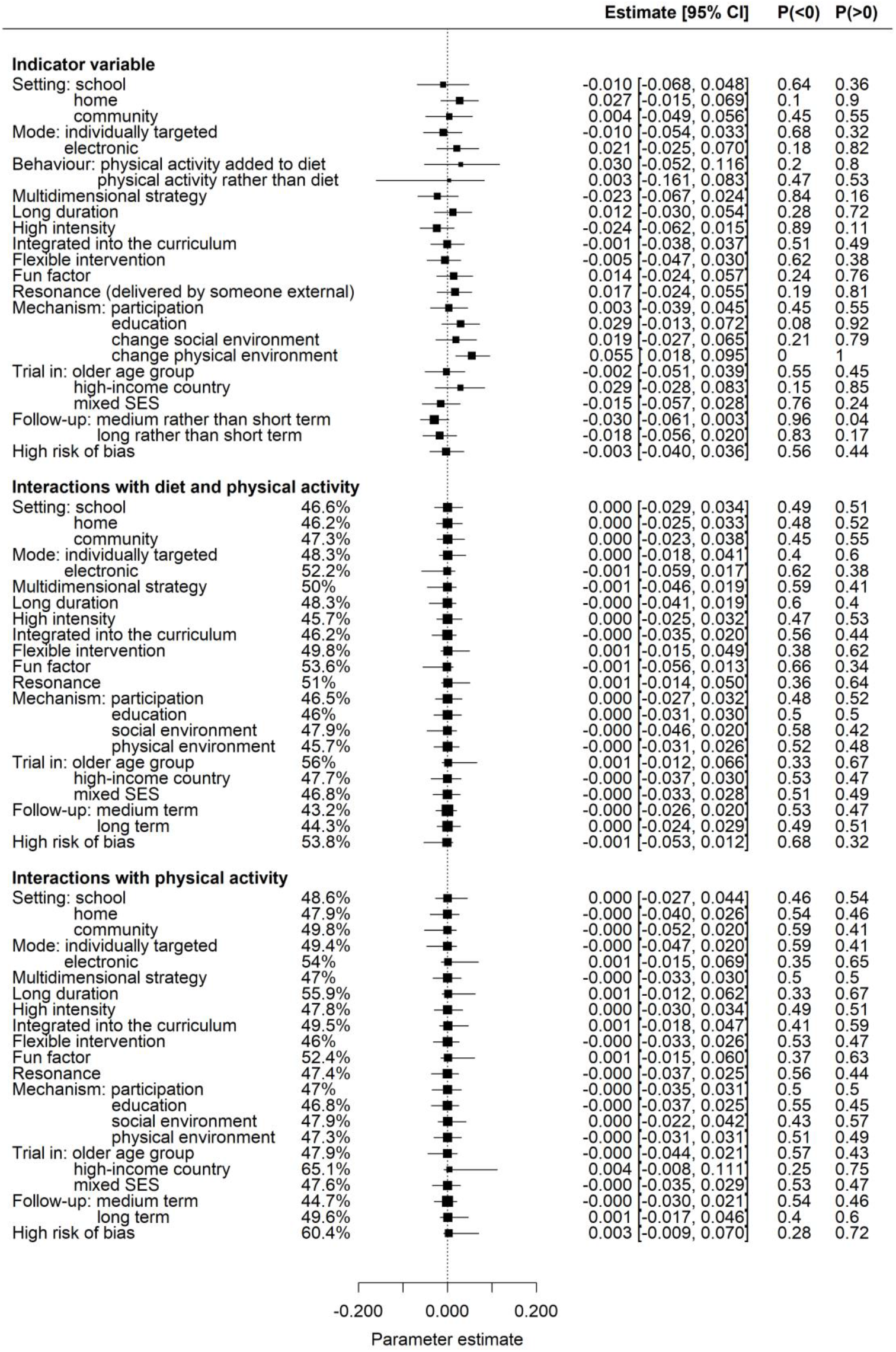
Parameter estimates from step 3 of the SSVS procedure (interactions between behaviour targeted (diet and/or activity) and all other indicators) for the primary (random effects) model. We list the probability that each coefficient is less than or greater than zero, *P*(< 0) and *P*(>0). For each interaction term we list the percentage of times it was selected by the SSVS model. The estimates of the intercept and heterogeneity parameter are *α* [95% *CI*] = −0.039 [−0.054, −0.023] and *τ* [95% *CI*] = 0.084 [0.071, 0.098] respectively.

#### I.2 Secondary analysis (fixed effects)

##### I.2.1 Step 1: No interactions

**Figure S6:**
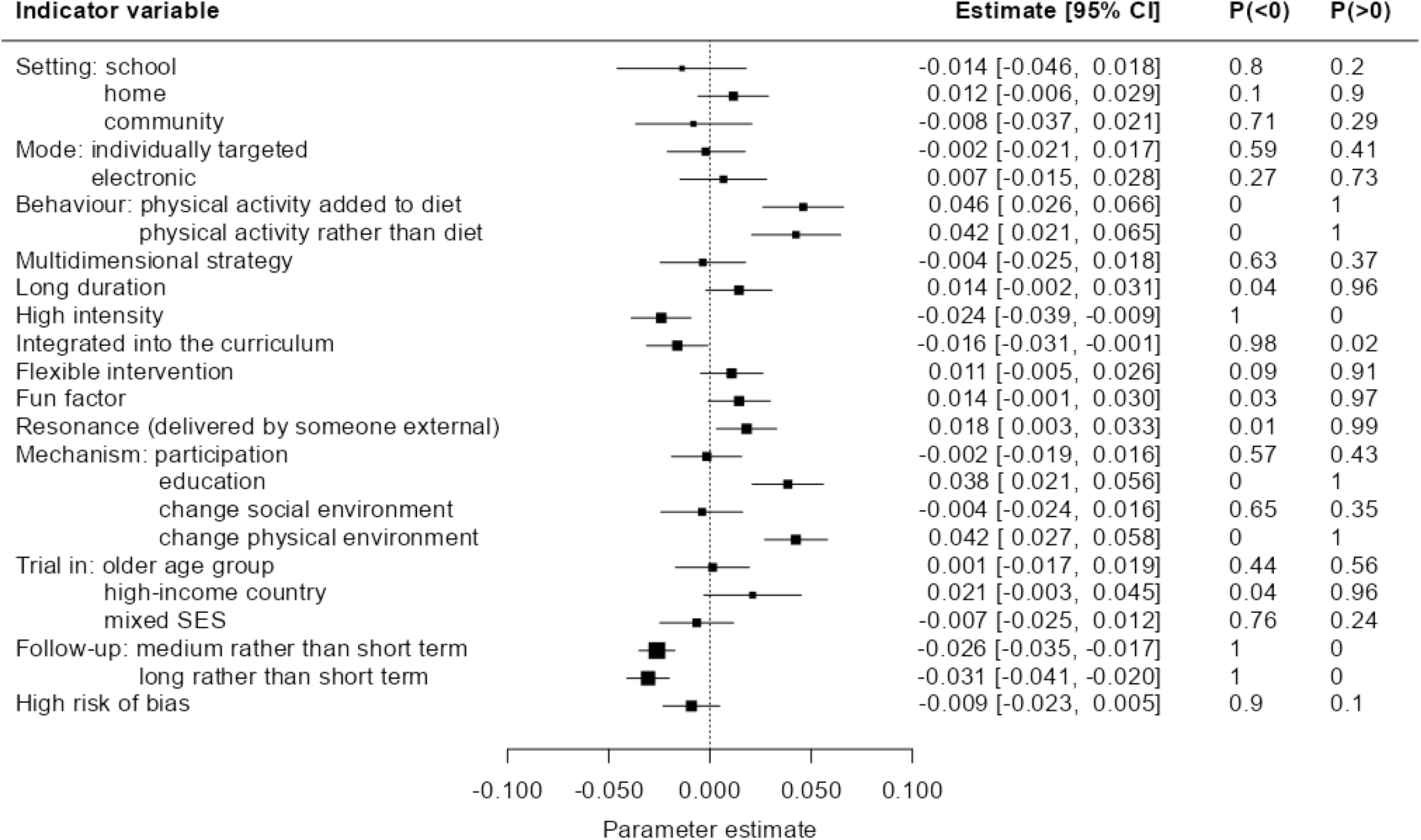
Parameter estimates from step 1 of the SSVS procedure (model with no interactions) for the secondary (fixed effects) model. We list the probability that each coefficient is less than or greater than zero, *P*(< 0) and *P*(>0). The estimate of the intercept is *α* [95% *CI*] = −0.033[−0.041, −0.025].

##### I.2.2 Step 2: Interactions with age

**Figure S7:**
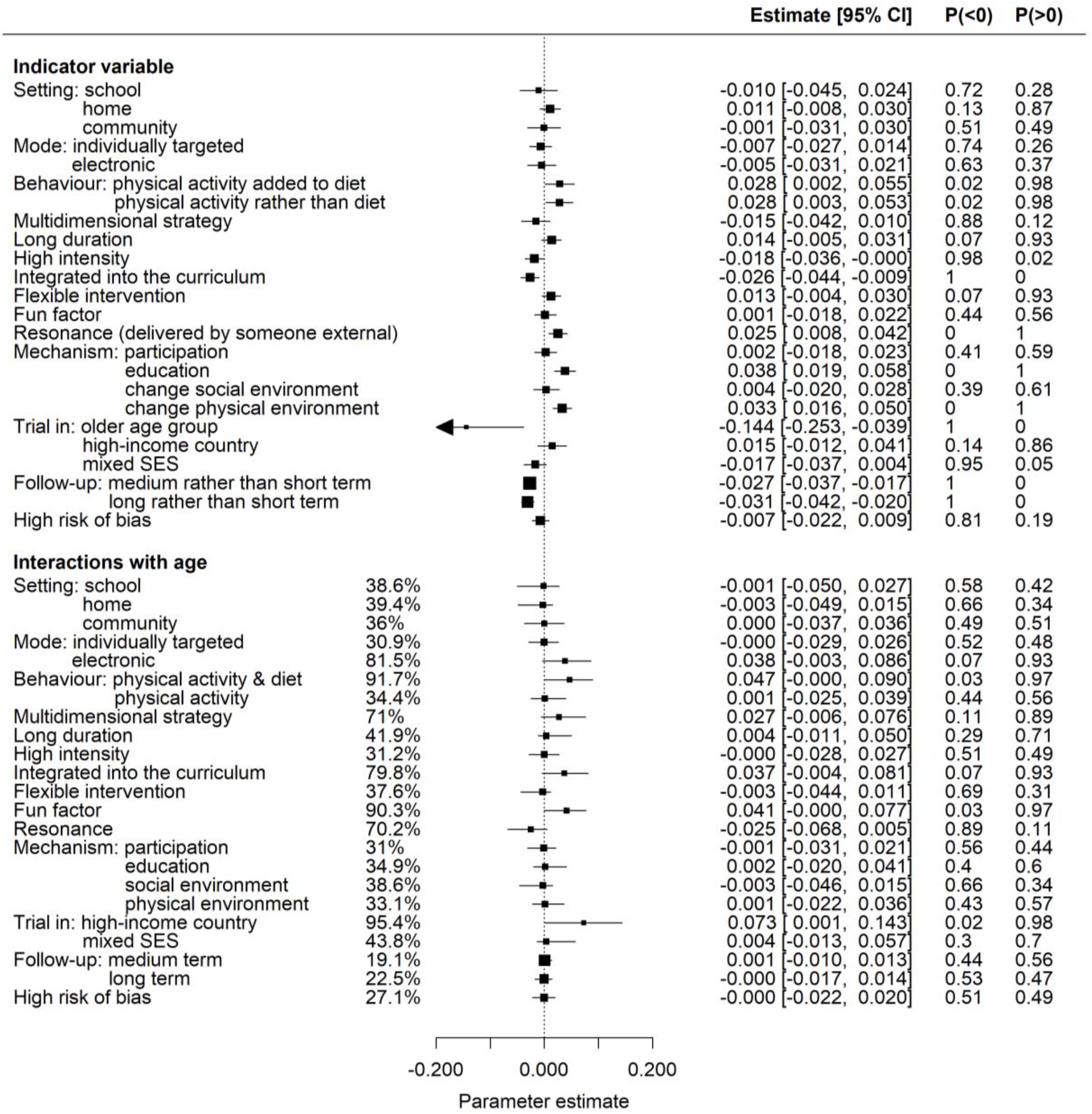
Parameter estimates from step 2 of the SSVS procedure (interactions between age and all other indicators) for the secondary (fixed effects) model. For each interaction term we list the percentage of times it was selected by the SSVS model. We list the probability that each coefficient is less than or greater than zero, *P*(< 0) and *P*(>0). The estimate of the intercept is *α* [95% *CI*] = −0.034 [−0.042, −0.026].

##### I.2.3 Step 3: interactions with change in behaviour targeted

**Figure S8:**
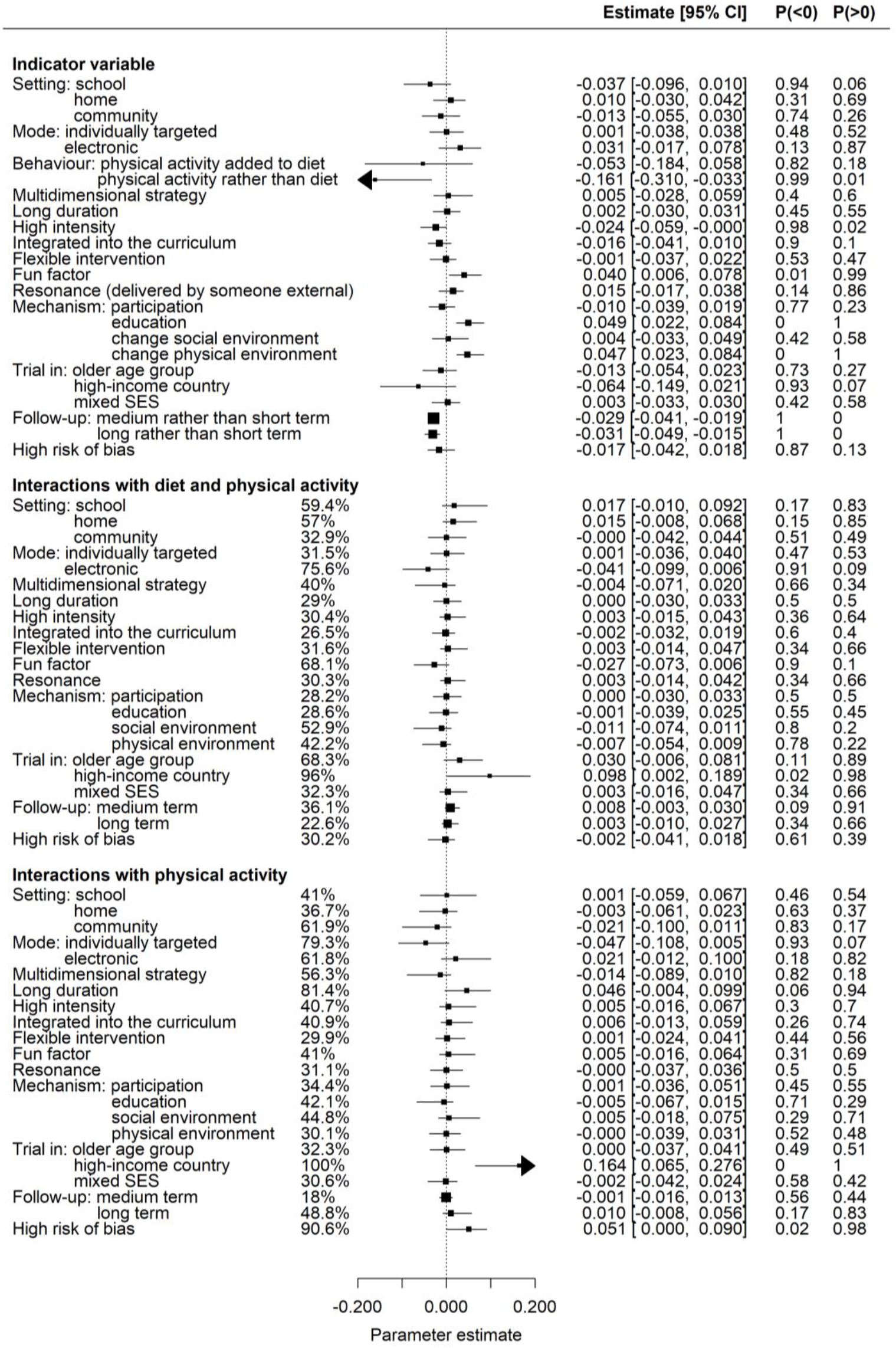
Parameter estimates from step 3 of the SSVS procedure (interactions between behaviour targeted (diet and/or activity) and all other indicators) for the secondary (fixed effects) model. We list the probability that each coefficient is less than or greater than zero, *P*(< 0) and *P*(>0). For each interaction term we list the percentage of times it was selected by the SSVS model. The estimate of the intercept is *α* [95% *CI*] = −0.032 [−0.041, −0.024].

### J Sensitivity analysis: separate analyses of different outcome scales

Here we show the results of the sensitivity analyses with different outcome scales analysed separately, i.e., excluding any mapped data. The analysis with zBMI outcomes alone is based on 171 observations from 110 trials while the BMI only analysis involves 182 observations from 129 trials. We also show the results with zBMI and mapped percentile (excluding mapped BMI and proportions) which includes an additional 25 observations from 18 trials compared with the zBMI only analysis.

On the whole, the results for BMI only are similar to the results of the primary analysis, while the zBMI only results are largely uncertain. The heterogeneity parameter is estimated to be smaller in the analyses of individual outcomes than in the full dataset. We summarize the comparisons between the results for the separate outcome scales in **Table S*3*** and **Table S*4***.

We begin by inspecting the effects with probability ≥0.95 in either the random-effects analysis or the fixed effects analysis and probability ≥0.9 in the other (those considered ‘robust’ in Section L below). There is strong evidence that environmental components are less beneficial in all three sensitivity analyses. The beneficial effects of physical activity alone and medium-term follow-up are also corroborated by the separate analyses, but with stronger evidence from BMI and weaker evidence (in the same direction) from zBMI. However, while there is no evidence of an effect of physical activity in the combined zBMI and percentile analysis, the direction is reversed. The observation that interventions are more effective in the older age group appears to be dominated by the BMI data (P(< 0) = 1), with no evidence of an age effect from zBMI only (P(< 0) = 0.31) or the combined analysis of zBMI and mapped percentile (P(< 0) = 0.23).

Of the effects with reasonable evidence (probabilities ≥0.9) in the primary analysis, the beneficial effect of integration and the less beneficial effect of home-based activities are supported by the zBMI alone and combined zBMI and percentile analyses, but with no evidence from BMI alone. The beneficial effect of multi-component strategies is in the same direction in all analyses with strong evidence from zBMI alone (P(< 0) = 0.96) and combined zBMI and percentile (P(< 0) = 0.95), and slightly weaker evidence from BMI only (P(< 0) = 0.83). The only conflicting evidence we observe is for the average effect of high-income countries: BMI strongly indicates a preference for high income countries (P(< 0) = 1), while zBMI finds greater beneficial effects for low income countries (P(>0) = 0.95). This may be due to a lack of data from low-income countries. The strength of this evidence for the zBMI analysis is increased with the addition of the mapped percentile data (P(>0) = 0.98).

For the interaction terms, we observe no conflicting evidence between the analyses on separate outcome scales. However, most of the effects observed in the primary analysis are dominated by one (or two) outcome scales with no evidence on the other(s). For example, the interactions between age and combined diet and physical activity, age and integration, age and income status of country, and activity and income status of country are observed in the BMI only analysis, but there is no evidence of these interactions from zBMI either alone or combined with percentile. Conversely, the interaction between activity and duration is observed in the same direction in all three analyses with the strongest evidence from zBMI and percentile (P(>0) = 0.78), followed by zBMI alone (P(>0) = 0.69) and BMI alone (P(>0) = 0.59).

The only interaction that appears with strong evidence in one of the separate outcome analyses that does not appear in the primary analysis is between interventions with an electronic component and interventions targeting both diet and physical activity. There is strong evidence of a synergistic effect in the BMI only analysis (P(< 0) = 0.97), and weaker evidence in the same direction for combined zBMI and percentile (P(< 0) = 0.89) and the zBMI only analysis (P(< 0) = 0.58).

#### J.1 zBMI only

**Figure S9:**
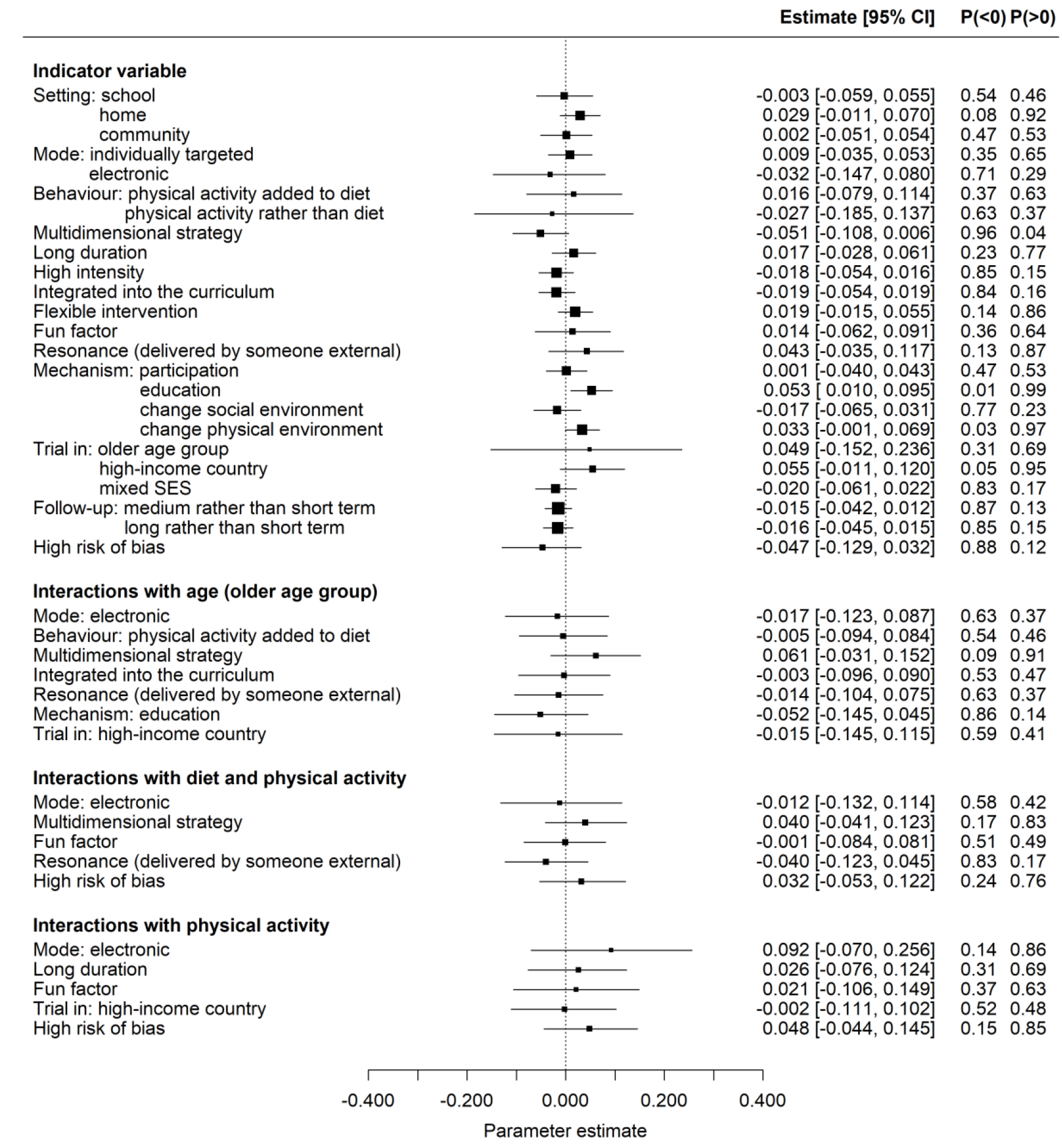
Parameter estimates from the random effects model fitted to data on reported zBMI only. The estimates of the intercept (for centred data) and heterogeneity parameter are *α* [95% *CI*] = −0.045[−0.061, −0.029] and *τ* [95% *CI*] = 0.049 [0.037, 0.063] respectively.

#### J.2 BMI only

**Figure S10:**
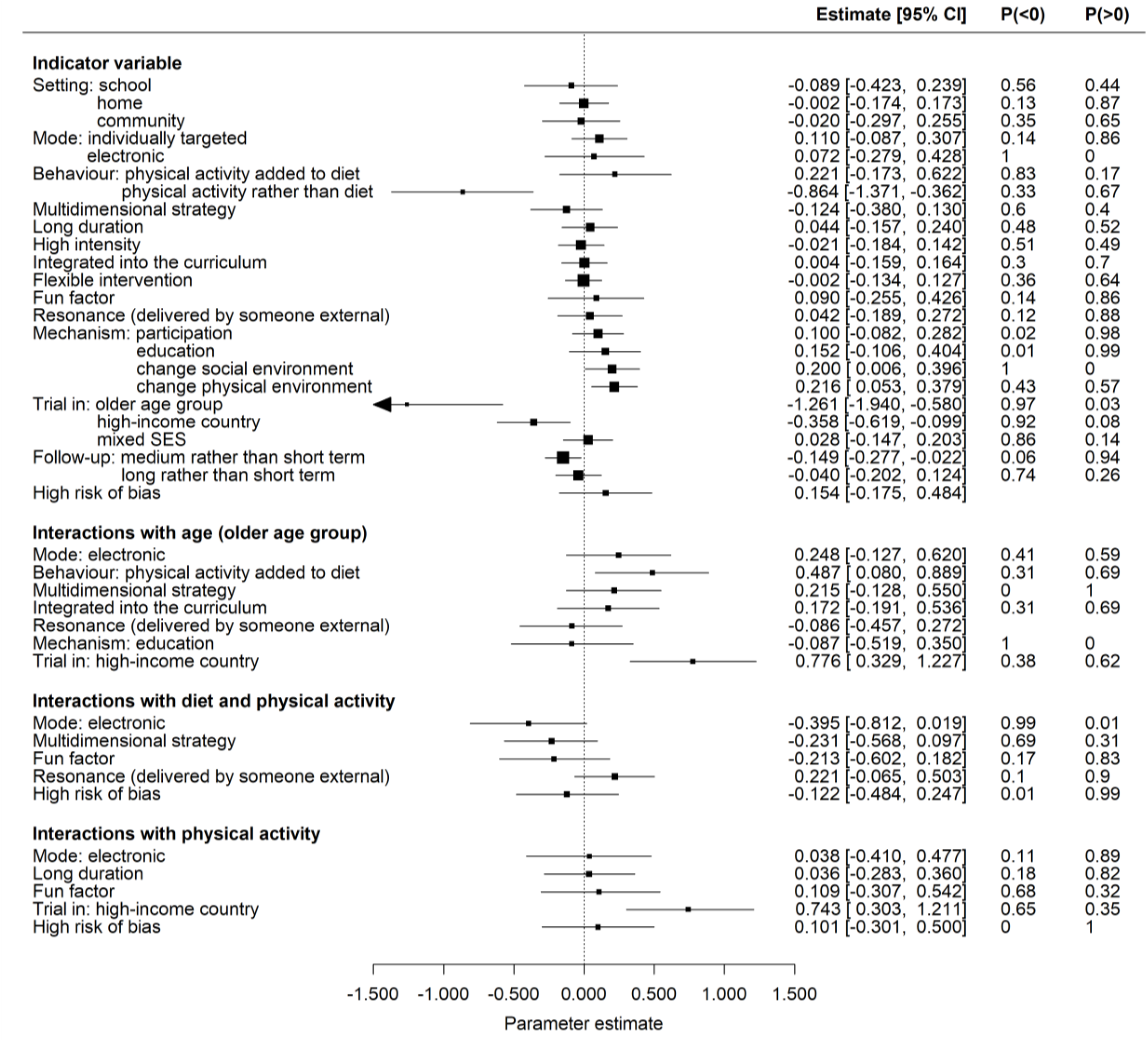
Parameter estimates from the random effects model fitted to data on reported zBMI only. The estimates of the intercept (for centred data) and heterogeneity parameter are *α* [95% *CI*] = −0.05 = −0.060[−0.133, −0.015] and *τ* [95% *CI*] = 0.271 [0.219, 0.332] respectively.

##### J.3 zBMI and percentile only

**Figure S11:**
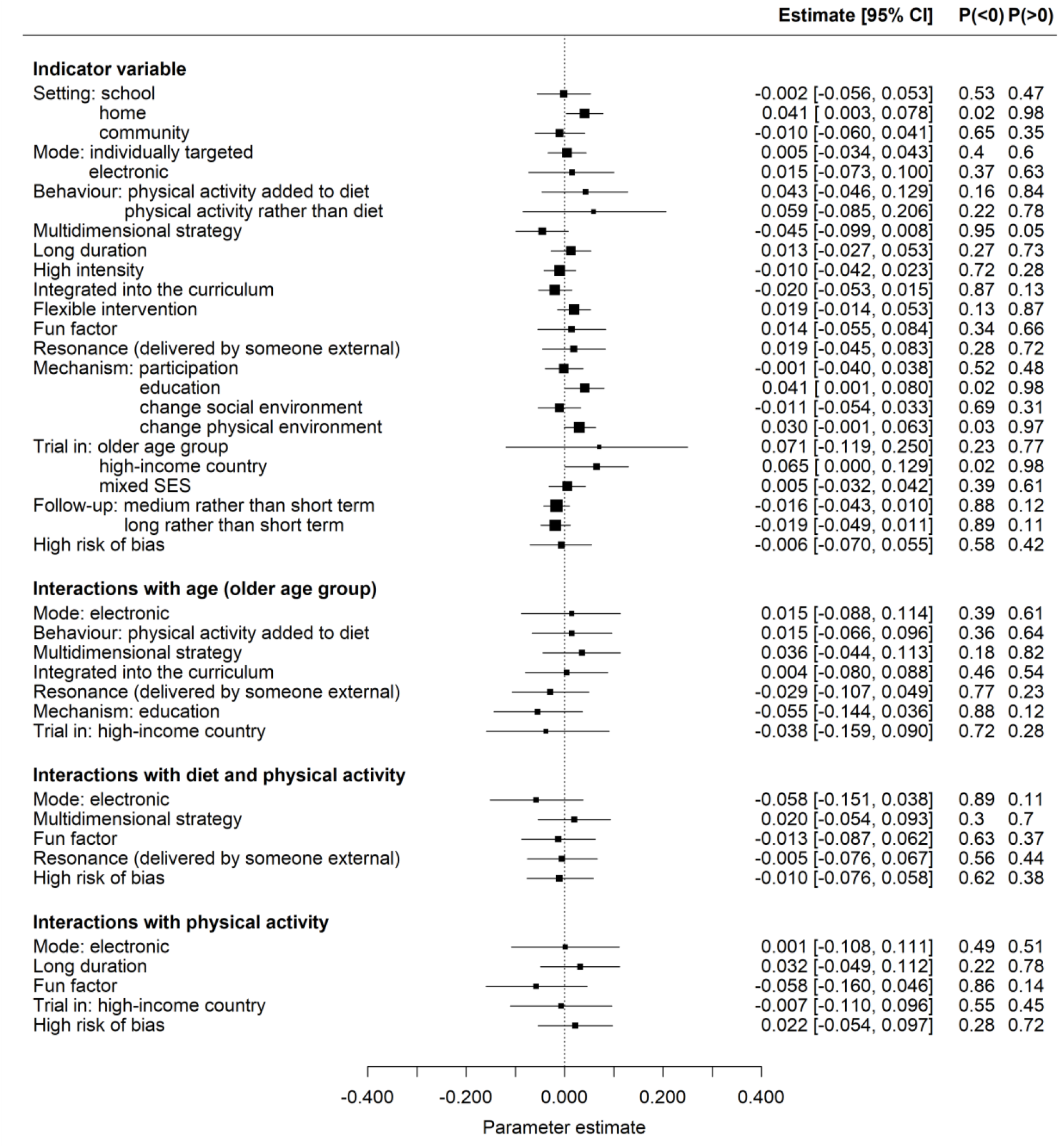
Parameter estimates from the random effects model fitted to data on reported zBMI and mapped percentile (excluding mapped BMI). The estimates of the intercept (for centred data) and heterogeneity parameter are *α* [95% *CI*] = −0.045[−0.060, −0.029] and *τ* [95% *CI*] = 0.051 [0.039, 0.065] respectively.

##### J.4 Comparison of different outcome scales

**Table S3:** A table summarizing the main differences between the indicator parameters estimated from the analyses on different outcome scales. The primary analysis includes all the reported zBMI data as well as the BMI and percentile data mapped onto the zBMI scale. We list the probability that a parameter estimate falls one side of zero in each analysis (with the corresponding direction indicated in the second column).

| Indicator | Direction of effect<br>(in main analysis) | Probabilities in different analyses |  |  |  |
| --- | --- | --- | --- | --- | --- |
|  |  | Primary | BMI only | zBMI only | zBMI and Percentile |
| Home | Less beneficial, $P(> 0)$ | 0.95 | 0.49 | 0.92 | 0.98 |
| Physical activity | Beneficial, $P(< 0)$ | 1 | 1 | 0.63 | 0.22 |
| Multi-strategy | Beneficial, $P(< 0)$ | 0.92 | 0.83 | 0.96 | 0.95 |
| Integrated | Beneficial, $P(< 0)$ | 0.93 | 0.48 | 0.84 | 0.87 |
| Social | Less beneficial, $P(> 0)$ | 0.77 | 0.98 | 0.23 | 0.31 |
| Environment | Less beneficial, $P(> 0)$ | 1 | 0.99 | 0.97 | 0.97 |
| Older age group | Beneficial, $P(< 0)$ | 1 | 1 | 0.31 | 0.23 |
| High income country | Beneficial, $P(< 0)$ | 0.95 | 1 | 0.05 | 0.02 |
| Medium term | Beneficial, $P(< 0)$ | 0.99 | 0.99 | 0.87 | 0.88 |

**Table S4:** A table summarizing the main differences between the interactions estimated from the analyses on different outcome scales. The primary analysis includes all the reported zBMI data as well as the BMI and percentile data mapped onto the zBMI scale. We list the probability that an interaction estimate falls one side of zero in each analysis (with the corresponding direction indicated in the second column).

| Interaction | Direction of effect<br>(in main analysis) | Probabilities in different analyses |  |  |  |
| --- | --- | --- | --- | --- | --- |
|  |  | Primary | BMI only | zBMI only | zBMI and Percentile |
| Older age & Diet and physical activity | Antagonistic, $P(> 0)$ | 0.99 | 0.99 | 0.46 | 0.64 |
| Older age & integration | Antagonistic, $P(> 0)$ | 0.94 | 0.82 | 0.47 | 0.54 |
| Older age & high income country | Antagonistic, $P(> 0)$ | 1 | 1 | 0.41 | 0.28 |
| Diet and physical activity & electronic | Synergistic, $P(< 0)$ | 0.84 | 0.97 | 0.58 | 0.89 |
| Physical activity & long duration | Antagonistic, $P(> 0)$ | 0.97 | 0.59 | 0.69 | 0.78 |
| Physical activity & high income country | Antagonistic, $P(> 0)$ | 1 | 1 | 0.48 | 0.45 |

### K Sensitivity analysis: assuming different correlations between repeated measures over time

Here we show the results of the random effects analysis assuming correlations of 0.5 and 0.95 between observations at different time points. We observe negligible differences from the primary analysis across all estimated effects. The estimate of the heterogeneity parameter is smaller for the 0.5 correlation (0.076 [0.063, 0.091]) and larger for the 0.95 correlation (0.088 [0.077, 0.010]).

#### K.1 Correlation of 0.5

**Figure S12:**
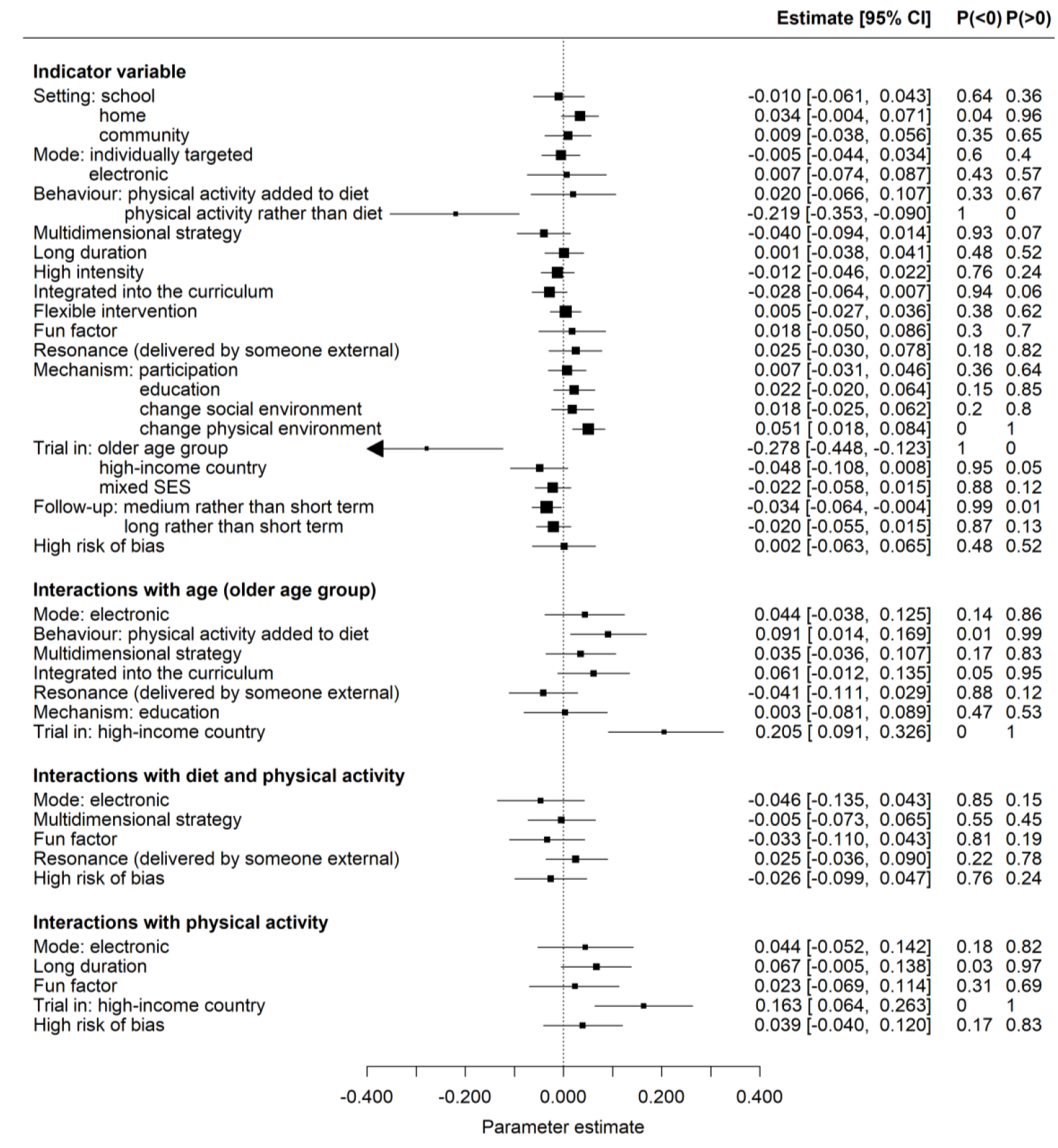
Parameter estimates from the random effects model assuming correlations of *ρ*_*y*,*tt*′_ = *ρ*_*d*,*tt*′_ = 0.5 for one degree of separation (short to medium, and medium to long). The estimates of the intercept (for centred data) and heterogeneity parameter are *α* [95% *CI*] = −0.037 [−0.052, −0.022] and *τ* [95% *CI*] = 0.076 [0.063, 0.091] respectively.

#### K.2 Correlation of 0.95

**Figure S13:**
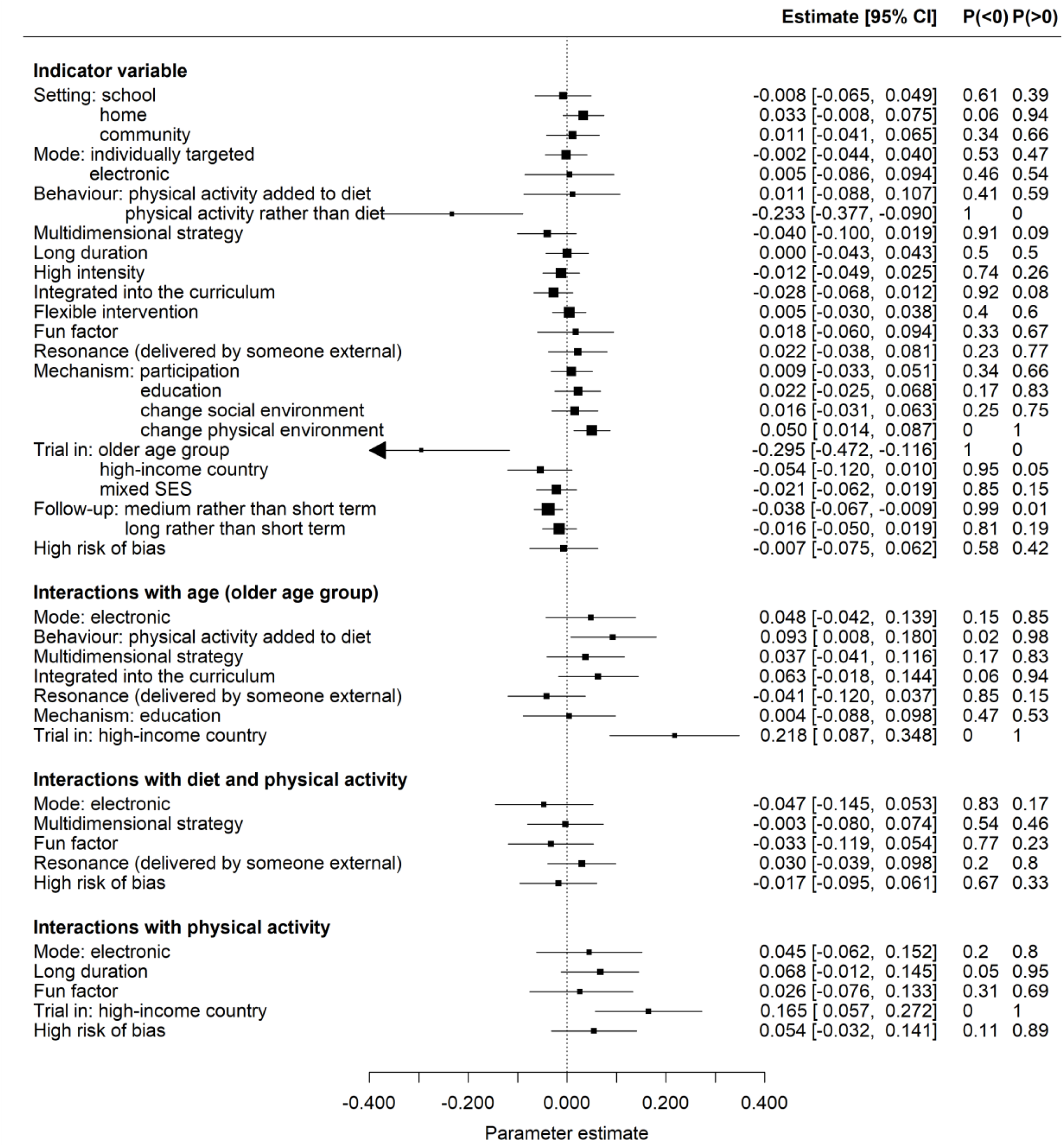
Parameter estimates from the random effects model assuming correlations of *ρ*_*y*,*tt*′_ = *ρ*_*d*,*tt*′_ = 0.95 for one degree of separation (short to medium, and medium to long). The estimates of the intercept (for centred data) and heterogeneity parameter are *α* [95% *CI*] = −0.037 [−0.053, −0.021] and *τ* [95% *CI*] = 0.088 [0.077, 0.010] respectively.

### L Comparing random and fixed effects models

Here we summarize the differences between the RE and FE analyses. We identify effects with strong evidence as those with probabilities in either direction (P(< 0) or P(>0)) that are greater than or equal to 0.95. For probabilities that exceed or equal 0.9, we label these effects as having reasonable evidence. Any effect which has strong evidence in the same direction in both analyses, we consider ‘robust’. Any effect which has strong evidence in one analysis and reasonable evidence in the other, we consider ‘almost robust’. Based on this procedure, we find robust indicator effects for physical activity alone versus diet alone (beneficial), environmental components (less beneficial), older age group (beneficial), high-income countries (beneficial) and medium verses short term (beneficial). The beneficial effect of integration is almost robust. We find robust antagonistic interactions between (i) the older age group and diet and physical activity interventions, (ii) the older age group and high-income countries, (iii) activity only and long interventions, and (iv) activity only interventions and high-income countries. Finally, we find an almost robust antagonistic interaction between the older age group and integrated interventions.

**Table S5:** A comparison of the direction and strength of evidence for indicator coefficients in the primary (random effects) and secondary (fixed effects) analyses. A coefficient direction <0 indicates a beneficial effect of an indicator coded as 1 and a direction >0 indicates a less beneficial effect. We categorize evidence as ‘strong’ if the probability that the coefficient takes a value one side of zero is greater than or equal to 0.95 and ‘reasonable’ if the probability is greater than or equal to 0.9. We label a coefficient as ‘robust’ if both analyses provide strong evidence in the same direction and ‘almost robust’ if one analyses is strong and the other is reasonable (in the same direction).

| Indicator | Primary analysis (RE) |  | Secondary analysis (FE) |  | Robustness |
| --- | --- | --- | --- | --- | --- |
|  | Direction | Strength | Direction | Strength |  |
| School | - | - | $<0$ | Strong | |
| Home | $>0$ | Strong | - | - | |
| Community | - | - | - | - |  |
| Individual | - | - | - | - |  |
| Electronic | - | - | - | - |  |
| Diet & activity | - | - | $<0$ | Strong | |
| Activity | $<0$ | Strong | $<0$ | Strong | Robust |
| Multi-strategy | $<0$ | Reasonable | - | - | |
| Duration | - | - | - | - |  |
| Intensity | - | - | $<0$ | Strong | |
| Integration | $<0$ | Reasonable | $<0$ | Strong | Almost robust |
| Flexibility/choice | - | - | - | - |  |
| Fun factor | - | - | $>0$ | Strong | |
| Resonance | - | - | $>0$ | Strong | |
| Participation | - | - | - | - |  |
| Education | - | - | $>0$ | Strong | |
| Social | - | - | - | - |  |
| Environment | $>0$ | Strong | $>0$ | Strong | Robust |
| Age | $<0$ | Strong | $<0$ | Strong | Robust |
| Income country | $<0$ | Strong | $<0$ | Strong | Robust |
| SES | - | - | - | - |  |
| Medium term | $<0$ | Strong | $<0$ | Strong | Robust |
| Long term | - | - | $<0$ | Strong | |
| Risk of bias | - | - | $<0$ | Strong | |

**Table S6:** A comparison of the direction and strength of evidence for interaction coefficients in the primary (random effects) and secondary (fixed effects) analyses. A coefficient direction <0 indicates a synergistic interaction between the two *indicator*s and a direction >0 indicates an antagonistic interaction. We categorize evidence as ‘strong’ if the probability that the coefficient takes a value one side of zero is greater than or equal to 0.95 and ‘reasonable’ if the probability is greater than or equal to 0.9. We label a coefficient as ‘robust’ if both analyses provide strong evidence in the same direction and ‘almost robust’ if one analyses is strong and the other is reasonable (in the same direction).

|  | Primary analysis (RE) |  | Secondary analysis (FE) |  | Robustness |
| --- | --- | --- | --- | --- | --- |
| Interaction | Direction | Strength | Direction | Strength |  |
| Interactions with age |  |  |  |  |  |
| Electronic | - | - | - | - |  |
| Diet & activity | >0 | Strong | >0 | Strong | Robust |
| Multi-strategy | - | - | - | - |  |
| Integration | >0 | Reasonable | >0 | Strong | Almost robust |
| Fun factor | Not selected |  | - | - |  |
| Resonance | - | - | <0 | Reasonable |  |
| Education | - | - | Not selected |  |  |
| Income country | >0 | Strong | >0 | Strong | Robust |
| Interactions with diet & activity |  |  |  |  |  |
| School | Not selected |  | >0 | Strong |  |
| Home | Not selected |  | >0 | Reasonable |  |
| Electronic | - | - | <0 | Strong |  |
| Multi-strategy | - | - | Not selected |  |  |
| Fun factor | - | - | <0 | Strong |  |
| Resonance |  |  | Not selected |  |  |
| Social | Not selected |  | - | - |  |
| Income country | Not selected |  | >0 | 1 |  |
| Risk of bias | - | - | Not selected |  |  |
| Interactions with activity |  |  |  |  |  |
| Community | Not selected |  | <0 | Strong |  |
| Individual | Not selected |  | <0 | Strong |  |
| Electronic | - | - | >0 | Reasonable |  |
| Multi-strategy | Not selected |  | <0 | Reasonable |  |
| Duration | >0 | Strong | >0 | Strong | Robust |
| Fun factor | - | - | Not selected |  |  |
| Income country | >0 | Strong | >0 | Strong | Robust |
| Risk of bias | - | - | >0 | Strong |  |

